# PRIMEtime: an epidemiological model for informing diet and obesity policy

**DOI:** 10.1101/2022.05.18.22275284

**Authors:** Linda J Cobiac, Cherry Law, Peter Scarborough

**Affiliations:** School of Medicine and Dentistry Griffith University; Department of Agri-Food Economics and Marketing University of Reading; Nuffield Department of Population Health University of Oxford

## Abstract

**Background:** Mathematical modelling can play a vital role in guiding public health action. In this paper, we provide an overview of the revised and updated PRIMEtime model, a tool for evaluating health and economic impacts of policies impacting on diet and obesity. We provide guidance on populating PRIMEtime with country-specific data; and illustrate its validation and implementation in evaluating a combination of interventions in the UK: a sugar-sweetened beverage (SSB) tax; a ban on television advertising of unhealthy foods; and a weight loss program.

**Methods:** PRIMEtime uses routinely available epidemiological data to simulate the effects of changes in diet and obesity on 19 non-communicable diseases, in open- or closed-population cohorts, over time horizons from 1 year to a lifetime. From these simulations, the model can estimate impact of a policy on population health (obesity prevalence, cases of disease averted, quality-adjusted life years), health and social care costs, and economic measures (net monetary benefit, cost-effectiveness ratios). We populated PRIMEtime with UK data and validated epidemiological predictions against two published data collections. We then evaluated three current obesity intervention policies based on estimates of effectiveness from published evaluation studies.

**Results:** There was considerable variation in the modelled impact of interventions on prevalence of obesity and subsequent changes in health and the need for health care: restrictions on TV advertising of unhealthy foods to children led to the largest reductions in obesity prevalence; but the SSB tax, which also targeted adults, had the biggest benefits in reducing obesity-related disease; and the weight loss program, while having very small impact on obesity prevalence at the population scale, had large and immediate benefits in improving health and reducing health sector spending. From a health sector perspective, the combination of interventions produced a favourable net monetary benefit of £31,400 (12,200 to 50,700) million. But the combined effect in reducing prevalence of overweight and obesity, was not estimated to reach more than 0.81 percentage points (95% uncertainty interval: 0.21 to 1.4) for males and 0.95 percentage points (0.24 to 1.7) for females by 2050.

**Conclusions:** Diet and obesity interventions have the potential to improve population health and reduce health sector spending both immediately and in the long-term. Models such as PRIMEtime can be used to evaluate the economic merits of intervention strategies and determine how best to combine interventions to achieve maximum population benefit. But with almost a third of children and two-thirds of adults currently overweight or obese, we need to broaden the application of public health models to evaluating the structural and systemic changes that are needed in our society to address the underlying drivers of the obesity epidemic.

## Introduction

Overweight and obesity have been recognised as a cause of poor health for over 2000 years.[1] But in the early times, it is likely the overall prevalence of obesity in the population was low; with malnutrition and underweight a greater cause of ill-health and premature loss of life. It is only in the last two decades that the number of overweight and obese adults worldwide have outnumbered those who are underweight.[2] Globally, 39% of adults and 18% of children and adolescents are now estimated to be overweight or obese.[3] This is associated with an excess 4.7 million deaths and 147.7 million disability-adjusted life years, with around 90% of this burden due to cardiovascular diseases, diabetes, related kidney diseases, and cancers.[4]

Like many high-income countries, the United Kingdom has been at the leading edge of the transition to obesity. By the time the World Health Organization recognised the growing prevalence of overweight and obesity as a ‘global epidemic’ in 1998, around 3 in 5 adults in England were already either overweight or obese.[5] While successive governments have attempted to address the problem over the last 30 years, including 14 strategy documents and 689 proposed policies and programs, efforts have so far failed to turn the tide.[6, 7] Although rapid upwards trends in obesity prevalence of the 1990s had flattened by the early 2000s, projection models suggest that prevalence of overweight and obesity will remain high until at least 2035, without more effective action.[8]

Mathematical modelling can play a vital role in guiding public health action,[9] as we have witnessed in the COVD-19 pandemic. While an epidemic of obesity may be slower-moving than an epidemic of an infectious disease, the informative value of modelling is no less important. Models can help us understand what the likely consequences of an intervention are, how confident we can be in the predicted outcomes, and what are the risks associated with the decision to act (or not act).[10] In the absence of modelling, decision-making is little more than blind guesswork; but modelling will only be of value if the model’s design, its inputs and outputs are clearly communicated, and there is transparency in the assumptions that have been made, whether explicit in model parameters or implicit in the structural design.[9, 10]

PRIMEtime is an epidemiological modelling tool that was developed in the UK for simulating diet and obesity interventions and scenarios. The model was initially developed in 2014 for examining impacts of changes in diet and obesity on population health, including non-communicable disease rates and life expectancy,[11] and this was later expanded to include impacts on quality of life and costs of health and social care.[12] The model has been used to examine potential impacts of a range of interventions and scenarios, including a targeted weight loss program, regulation of television advertising of unhealthy foods to children, salt reduction, evaluating changes to dietary guidelines and identifying most environmentally sustainable dietary patterns.[11–16]

In recent years, we have made a number of updates to the PRIMEtime model. These include new features to capture background trends in obesity and disease rates, as well as updates of model parameters to incorporate the latest evidence on risks of disease associated with diet and obesity, and to capture the latest data on risk factor and disease rates in the UK. In addition, we have modified the cohort simulation to include options for both open- and closed-cohort simulation. While previously a closed-cohort simulation model, allowing users to examine changing health and cost consequences for an adult population cohort ageing through time, PRIMEtime now additionally incorporates open-cohort simulation of outcomes for children and future birth cohorts out to 2050. This feature enables users to examine impacts of interventions on population prevalence of overweight and obesity and population rates of disease into the future, and potentially facilitates linkage of PRIMEtime with economic and environmental simulation models.

In this paper we describe the updated PRIMEtime model and illustrate its use in evaluating a combination of interventions: a sugar-sweetened beverage tax; a ban on television advertising of unhealthy foods; and a weight loss program. These are all interventions that have been implemented or recommended in the government’s latest obesity strategy.[17] This work is not intended to be a comprehensive evaluation of all potential obesity intervention strategies, rather the aim is to provide transparency on the model’s design and input parameters, to illustrate how the model can be used, and discuss how this and other modelling may be developed and better integrated into policy in the future.

## Methods

### The PRIMEtime model

#### Model Design

The PRIMEtime modelling approach (Figure 1) consists of three interlinked components: (1) a risk factor exposure module, which quantifies the effect of change in prevalence of one or more risk factors on incidence of related diseases; (2) a series of disease models, which simulate changing incidence, prevalence and mortality of diseases related to the modelled risk factors; (3) a lifetable, which simulates overall impacts on the population, health and social care system, due to the changing disease epidemiology.

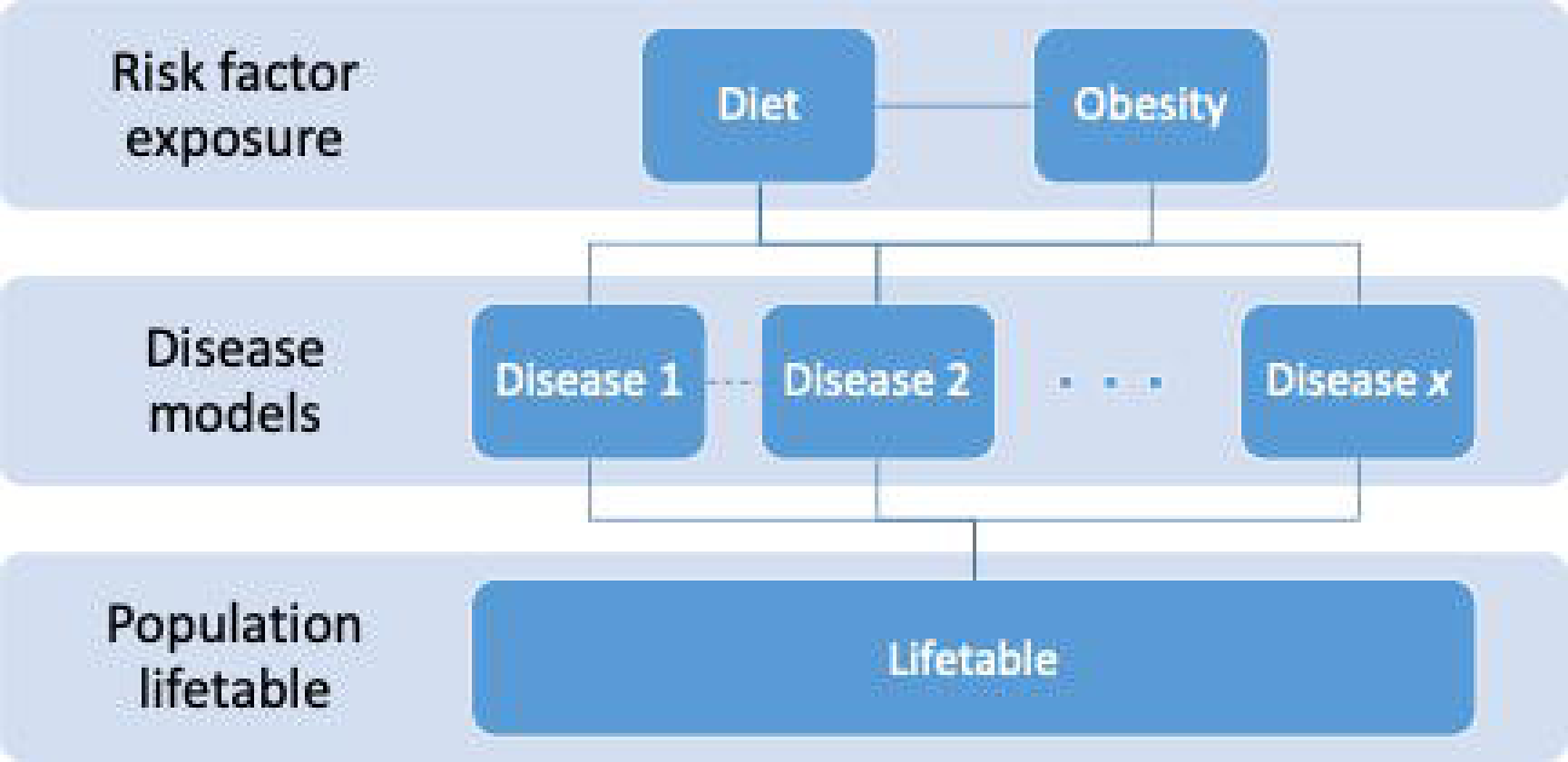

##### Risk factor exposure

When an intervention is simulated in PRIMEtime, the magnitude of impact on disease associated with a risk factor (e.g. high body mass index) depends on a combination of variables: (1) the baseline population exposure to the risk factor; (2) the measured effect of the intervention on exposure, and the proportion of the population who stand to benefit; and (3) the magnitude of the relative risk of disease associated with the risk factor. This impact is quantified by the population impact fraction (PIF):[18]

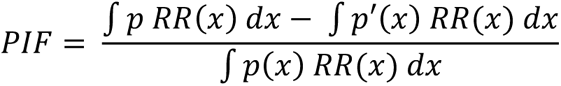

where: *x* is the units of a risk factor (e.g. body mass index); *p*(*x*) is the prevalence of that risk factor; *p*′(*x*) is the prevalence of the risk factor after a scenario or intervention; and *RR*(*x*) is the relative risk of disease associated with the risk factor. The PIF is age- and sex-dependent. Where there are background trends in the risk factor (e.g. obesity), the PIF is also time-dependent. Where diseases are influenced by multiple independent risk factors, PRIMEtime combines the PIFs multiplicatively:

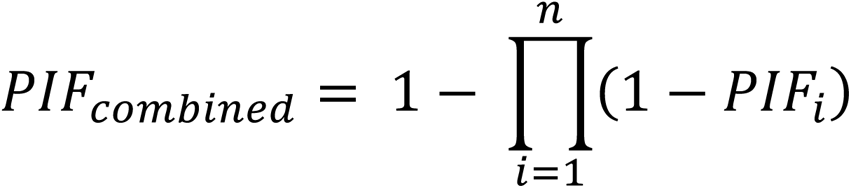

for *i* = 1…*n* diseases.

The relative risks of disease associated with each risk factor in PRIMEtime, have been drawn from systematic reviews and meta-analyses. For modelling purposes, it is necessary to establish dose-response relationships across the entire distribution of risk factor exposure in the population of interest. Large, long-term randomised controlled trials are rare, and where they do exist, they provide only limited variance between control and intervention arms in the risk factor of interest. Therefore, establishing causality in the relationship between a risk factor and condition relies heavily on prospective cohort studies, and evaluation of the entirety of evidence against criteria, such as those of Bradford-Hill – strength and consistency of evidence, specificity, temporality, biological gradient, plausibility, coherence, analogy and supporting experimental evidence.[19] In developing and updating PRIMEtime we have preferentially drawn on systematic reviews and meta-analyses that take these factors into account.

##### Disease models

Diet- and obesity-related diseases currently included in PRIMEtime include ischaemic heart disease, ischaemic stroke, intracerebral haemorrhage, hypertensive heart disease, diabetes mellitus type 2, colorectal cancer, breast cancer, uterine cancer, oesophageal cancer, kidney cancer, pancreatic cancer, liver cancer, multiple myeloma, asthma, low back pain, osteoarthritis of the hip and knee, depression, atrial fibrillation/flutter, and gallbladder/biliary diseases.

Cardiovascular diseases, diabetes and cancers are modelled in a three-state Markov model, in which the population is either in a healthy state, a diseased state or dead (from the disease or from other un-related causes).[20] The initial distribution of the population between the healthy and diseased states is determined by the prevalence of the disease in the baseline year. Further transition between states over time is based on annual rates of incidence, remission and case fatality. However, for the remaining PRIMEtime diseases that are chiefly characterised by morbidity rather than mortality (low back pain, osteoarthritis, depression, and gallbladder and biliary diseases) we do not model progression to mortality.

PRIMEtime is designed to evaluate the effects of interventions, scenarios or system shocks in *preventing* disease and the consequent effects on population health and society. Therefore, we model the effects of a change in risk factor exposure, which is quantified by the PIF, as a change in disease incidence rate (*I*):

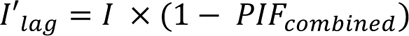

The calculation of *I*′*_lag_* allows for a time lag in the reversal of disease risk. While there is an absence of direct evidence examining the time course for changes in disease rates following population changes in dietary intake or obesity, the World Health Organization has estimated it takes three to five years to achieve full reversal of stroke risk and around two-thirds of heart disease risk, from trials of blood pressure treatments.[21] Risk reversibility of cancers is likely to be longer, based on follow-up of patients around ten years after bariatric surgery or intentional weight loss.[22]

##### Population lifetable

In the population lifetable component of PRIMEtime, the population is divided into five-year age and sex cohorts that are simulated through time. In a closed cohort simulation, the model is typically run until everyone in the starting population is dead. In an open cohort simulation, which incorporates future birth cohorts, the model can be run until the desired time horizon is reached or until there are no further birth cohorts entering the simulation.

Each population age- and sex-cohort is simulated in a proportional multi-state lifetable.[23] This consists of a standard lifetable model, which simulates the survival experience of a cohort based on age- and sex-specific mortality rates, connected with the series of disease-specific models described above. There are three key actions that occur in the proportional multi-state lifetable model.

First, when we simulate a change in prevalence of obesity, this influences incidence of obesity-related diseases, which, over time, lead to changes in disease-specific mortality rates. These changes in disease-specific mortality rates modify the total mortality rate in the main lifetable, while holding the proportion of mortality due to all other causes (i.e. causes not related to obesity) constant.

Second, everyone in the lifetable cohort who is alive has a utility weight, which reflects average ‘quality of life’ at each age and sex. In the same way that total mortality in the lifetable is modified by changes in disease-specific mortality, the total utility in the lifetable is modified by changes in disease-specific utility, which are derived in the disease-specific models from the change in incidence and/or prevalence of the disease and the disease-specific utility weight(s).

Third, everyone in the lifetable cohort who is alive has an average cost associated with their health care. This cost increases or decreases, based on changes in rates of obesity-related diseases, which are derived in the disease-specific models from the change in incidence and/or prevalence of the disease and the disease-specific health care cost.

Together, the lifetable, disease-specific and risk factor exposure models simulate each cohort ageing, over time, until everyone is dead, has reached 100 years of age, or the simulation has reached the end of its time horizon. From these simulations, PRIMEtime can determine the effect of a change in prevalence of a risk factor on: (a) population life expectancy; (b) gain or loss of quality-adjusted life years (QALYs); and (c) impact on costs to the health or social care system.

#### Country-specific input data

##### Population and mortality

PRIMEtime can be adapted for any country context with a number of country-specific inputs. Population is entered by sex and 5-year age group, with the exception of the 0 to 4 year old group, which is split into 0 year olds and 1 to 4 years olds to facilitate calculation of life expectancy at birth. To run a closed cohort analysis, only the population in the baseline year of analysis is required. If running an open cohort analysis, future cohorts of 0 to 4 years olds can be estimated from population projections. The model currently has capacity to accept new entrants into the modelling until 2050. PRIMEtime also requires mortality rates from all causes by sex and single year of age, from age 0 to 100+.

Both mortality rates and population numbers can be downloaded in the required format from the Human Mortality Database[24] for 41 countries and, in some cases, also by population sub-groups, e.g. by ethnicity. The Institute of Health Metrics Results Tool[25] also provides links to their collations of population, mortality and life table data for Global Burden of Disease analyses, which is an alternative source of data for countries not included in the Human Mortality Database or if within-country data collections are not accessible. These sources currently provide data up to 2019. Population estimates for later years (e.g. for open cohort analyses incorporating future birth cohorts) are based on population projections. The United Nations provide a database of global population projections if local estimates are not available.

##### Disease epidemiology

Population-specific disease rates can be derived from a variety of sources such as death and disease registries and disease-specific studies. Alternatively, PRIMEtime users can draw on Global Burden of Disease (GBD) syntheses of these sources. Cause-specific incidence, prevalence and mortality rates can be downloaded, by age and sex, using the GBD Results Tool.[25] These data can be used in PRIMEtime for non-commercial purposes under the Creative Commons Attribution-Non Commercial 4.0 International License.[26]

A key challenge in populating the PRIMEtime model is the requirement for cause-specific case fatality rates. In the PRIMEtime model, the case fatality reflects the excess mortality of someone with the disease compared with someone who does not have the disease. These rates are rarely reported, however they can be derived from cause-specific incidence, prevalence and mortality rates, together with population numbers and all-cause mortality rates, using the *disbayes* R package, which can be found on the GitHub repository.[27] This package is recommended as, using a Bayesian approach, it estimates case fatality using the same three-state model and assumptions that underpin disease simulation in the PRIMEtime model, so consistency in approach is maintained.

Background trends in cause-specific incidence and case fatality rates can be estimated by fitting models to rates derived over multiple years. The GBD Results Tool[25] contains yearly estimates of rates from 1990.

##### Risk factor exposure

Prevalence of risk factors can be derived from population survey data. PRIMEtime is designed to take the mean and standard deviation of the distribution of risk in the population, by age and sex. This allows the population impact fraction to be calculated continuously (e.g. where the relative risk is defined per unit of BMI) or categorically (e.g. where relative risks are defined for overweight and obese categories of prevalence). There is also an option for taking background trends in risk factors into account.

Derivation of risk factor prevalence and trends depend on the availability of survey data, which is highly country-specific. Cobiac et al[8] provides an example of deriving trends in body mass index from health survey data in England. However, some global collations of data also exist, such as the World Health Organization Global Health Observatory[3] and the Global Dietary Database.[28]

##### Costs

There is currently no global database of age- and sex-specific costs of disease treatment in different countries. For PRIMEtime analyses in the UK, the unit costs of treatment for the explicitly modelled diseases and the average costs associated with health care for all other diseases, have been derived from a range of administrative datasets. Briggs et al.[29] have published a detailed description of these methods.

#### Model Implementation

##### Simulation options

PRIMEtime provides a number of options for evaluation. The user can:

‒ define discount rates for health and cost impacts;
‒ define time horizons for population simulation and for output of data;
‒ set time lags on the effects of a risk factor on diseases;
‒ include or exclude background trends in risk factors;
‒ include or exclude diseases; and
‒ select population cohorts for simulation (a closed cohort analysis only simulates outcomes for the population alive in the baseline year of analysis, whereas an open cohort analysis also simulates outcomes for those born in future years).

These choices depend on the purpose of the PRIMEtime simulation. For example, calculating cost-effectiveness or return-on-investment of an intervention, will generally require discounting of cost and health outcomes. Comparing cost-effectiveness of multiple preventive interventions can generally be performed using a closed cohort simulation, since the relative performance of different interventions is unlikely to be greatly impacted by the addition of future birth cohorts who will not experience health benefits until well into the future. However, an open cohort simulation may be preferred if PRIMEtime is being used to simulate impacts of a scenario on rates of obesity-related diseases or costs of treating these diseases, over time into the future.

##### Scenario specification

PRIMEtime currently allows for simultaneous simulation of up to five scenarios or population sub-groups. While the model can be run any number of times for different evaluations, simultaneous simulation facilitates analyses where uncertainty in the relative order of or difference between scenarios or sub-groups is important. For example, by simultaneously simulating outcome by quintile of Index of Multiple Deprivation, it is possible to estimate uncertainty in the slope index of inequality, a measure of health inequality.

##### Probabilistic Sensitivity Analysis

The PRIMEtime model is designed to be run iteratively. If only one iteration is performed, the model will produce point-estimates of outputs based on the mean values of model input parameters. However, when run many times, the model will randomly select from uncertain distributions around input parameters to produce a range of outputs, from which mean or median values and 95% uncertainty intervals can be determined. The degree of uncertainty in outputs depends on the number of inputs that are uncertain, and the distribution of uncertainty around those inputs. As a rule-of-thumb, we recommend running as many iterations as are required to achieve stability in outputs to the desired number of significant figures. Typically, this is between 2000 and 5000 iterations.

### PRIMEtime evaluation of obesity interventions

#### Intervention specification

We demonstrate use of the PRIMEtime model in evaluating three interventions for addressing obesity in the UK population:

1. Implementing a Soft Drinks Industry Levy (SSB tax)
2. Banning television advertising of foods and drinks high in fat, sugar, and salt (HFSS) between 5:30 AM and 9:00 PM (TV ad bans).
3. Offering total diet replacement to adults (18+ years) with a BMI greater than 30 kg/m^2^ (Weight loss program).

Intervention effects were derived from previously published analyses as described in Table 1. For this study, we evaluated the interventions individually and additionally evaluated a combination of all three interventions, as if rolled out in the UK population from a baseline year of 2015.

**Table 1.**
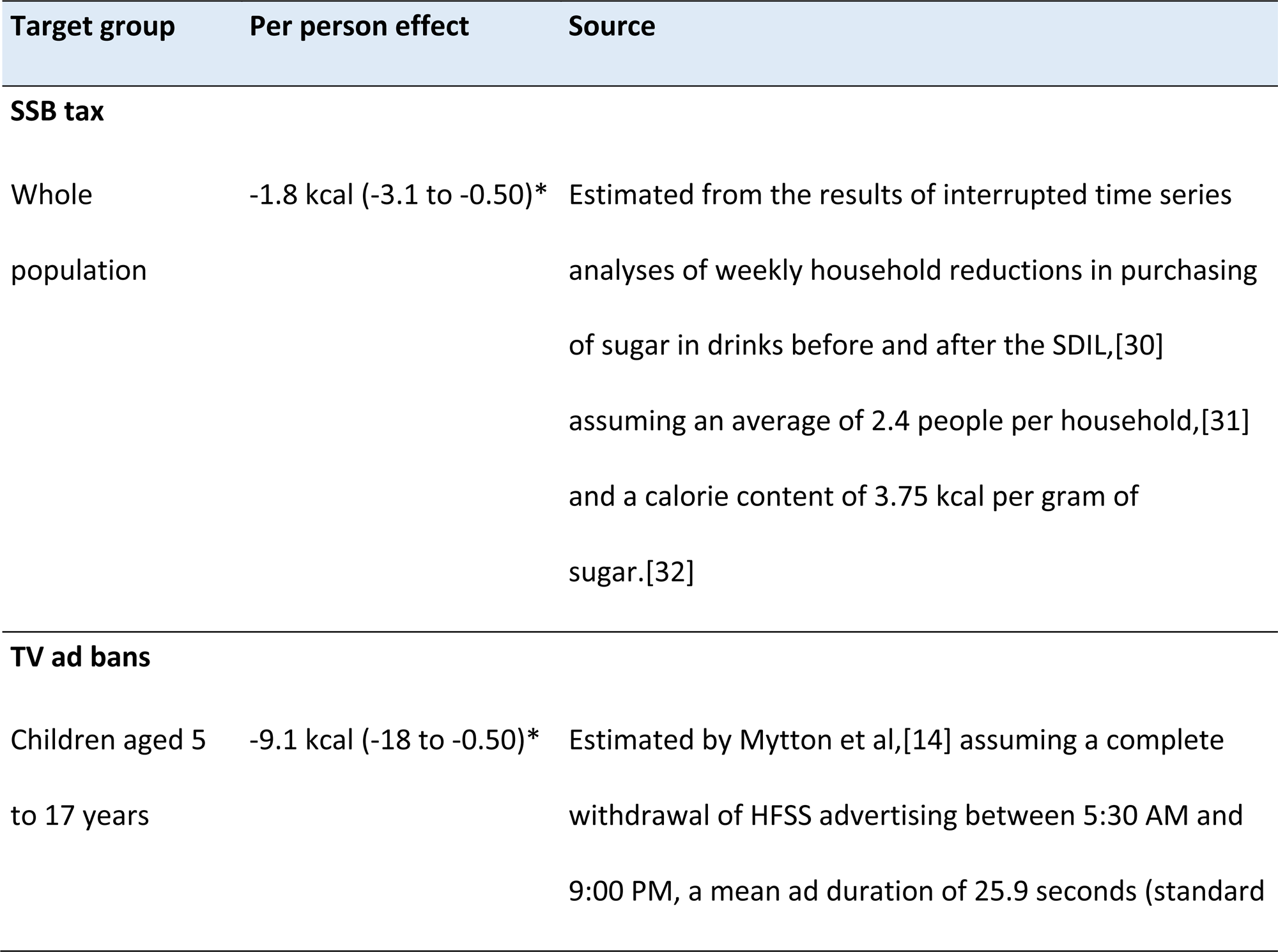
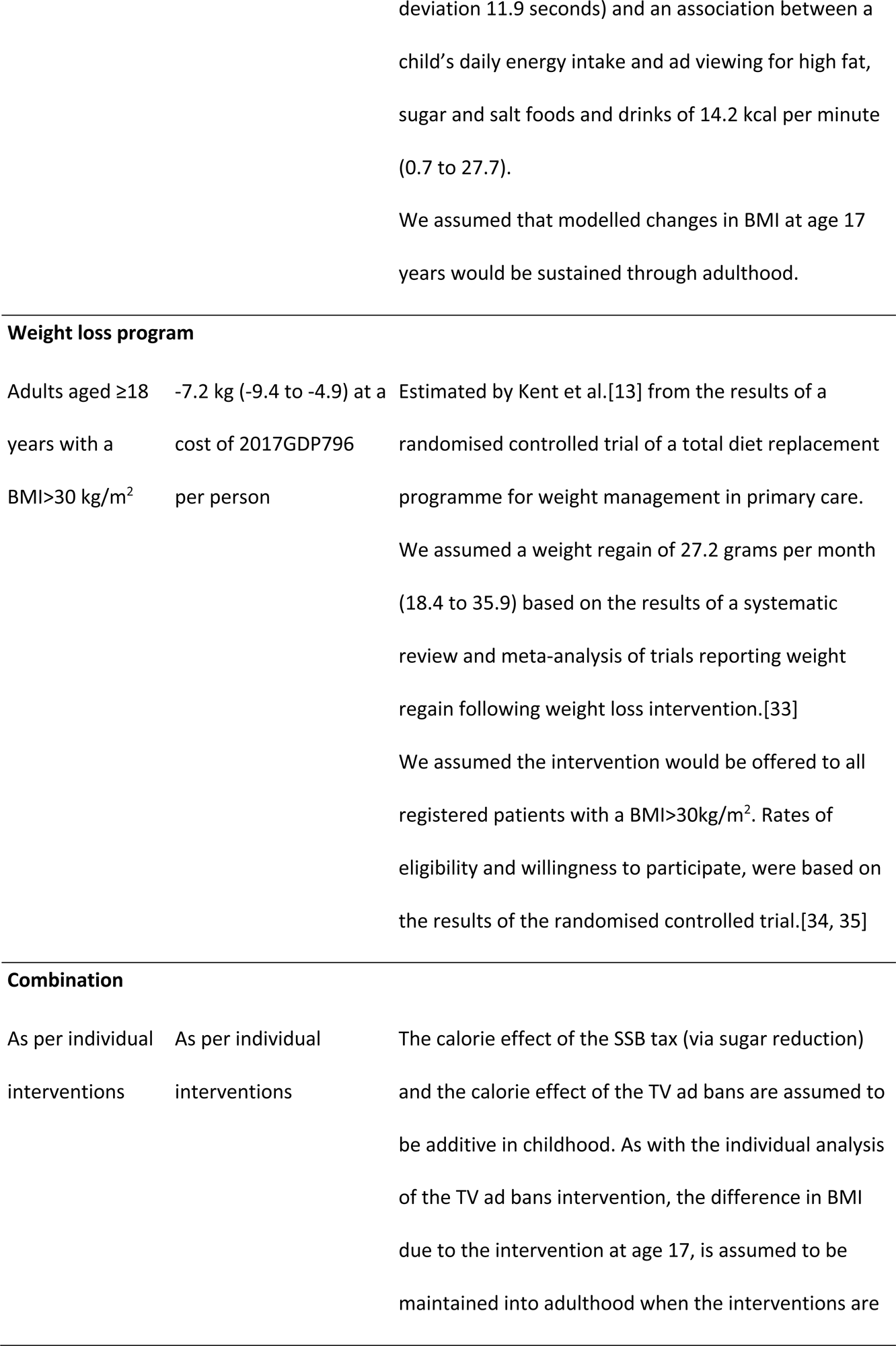
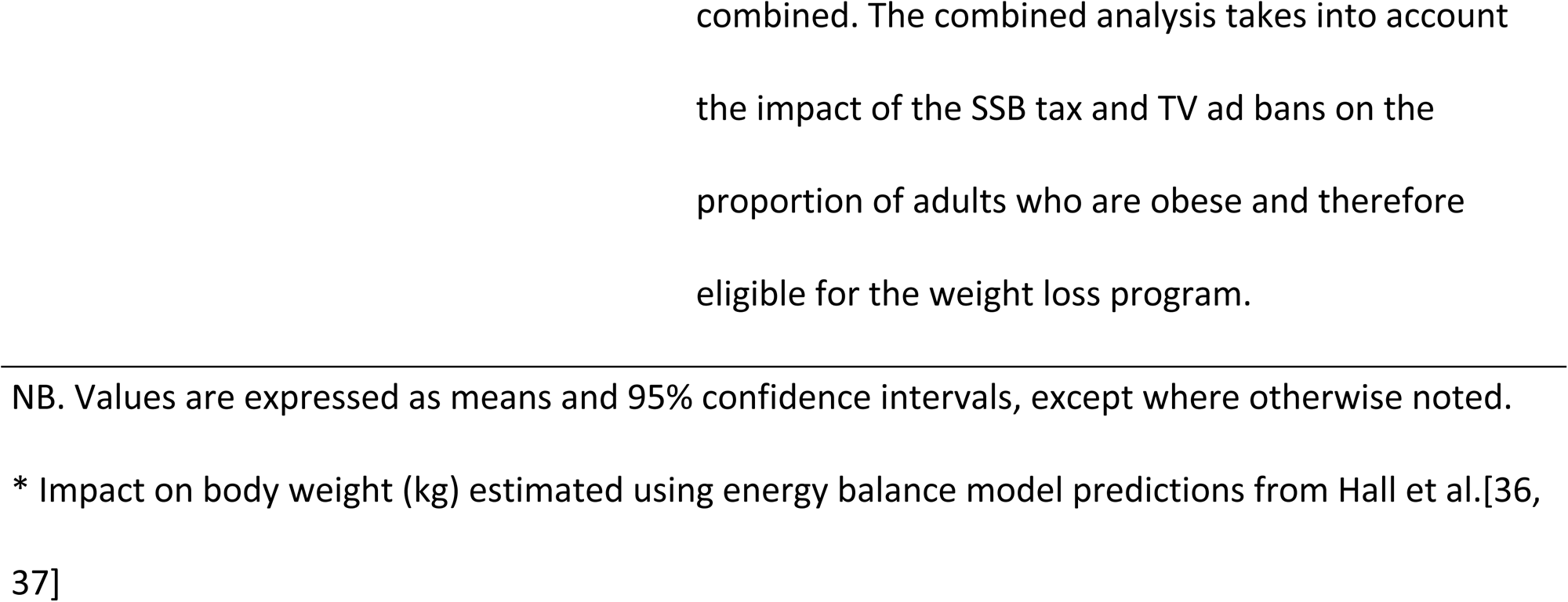
Intervention scenarios.

#### UK data inputs

For the obesity intervention evaluation, we populated the PRIMEtime model with data for the UK. Table 2 described the data sources and assumptions made, and Text S1 provides the data as entered into PRIMEtime.

**Table 2.**
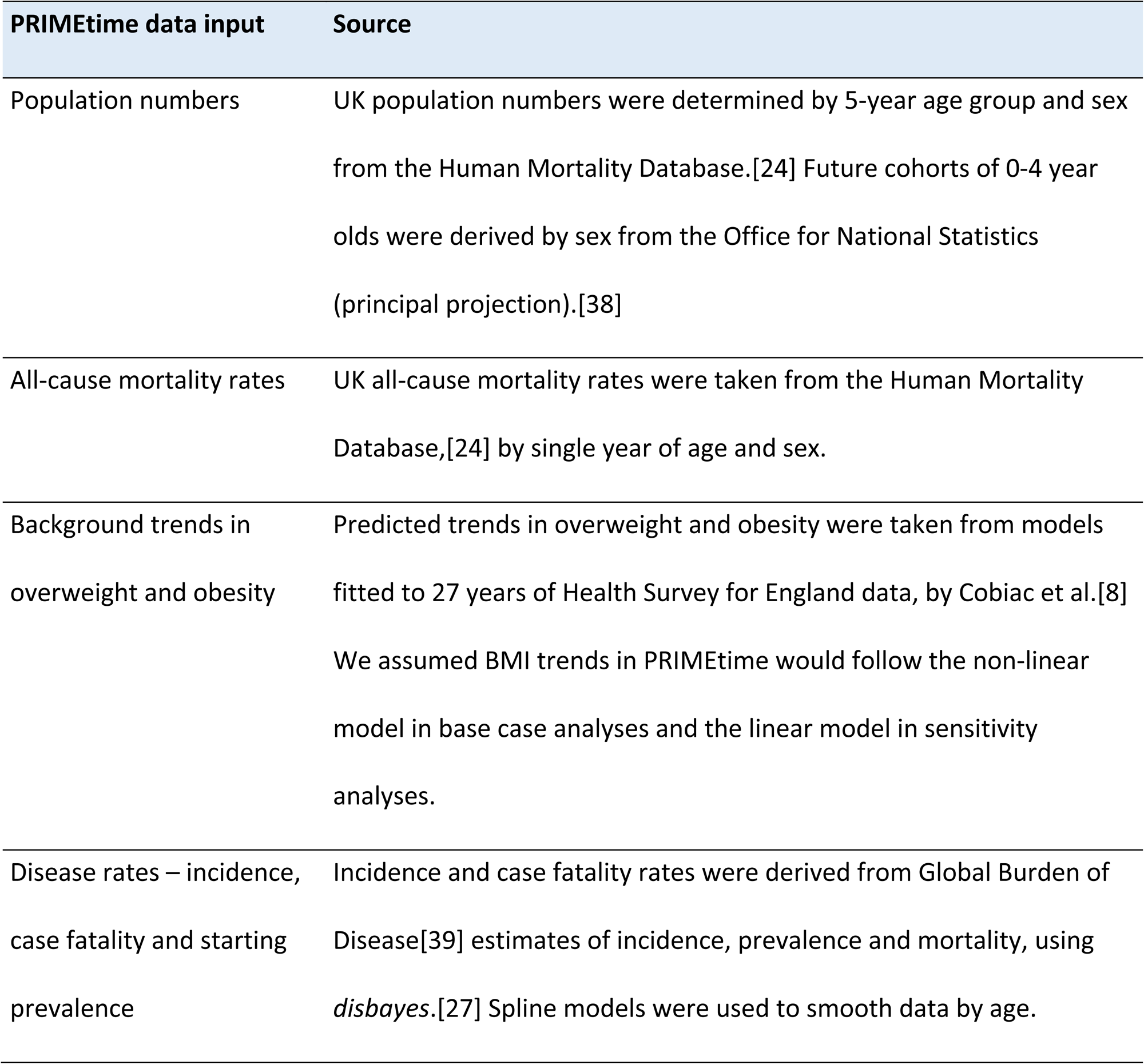
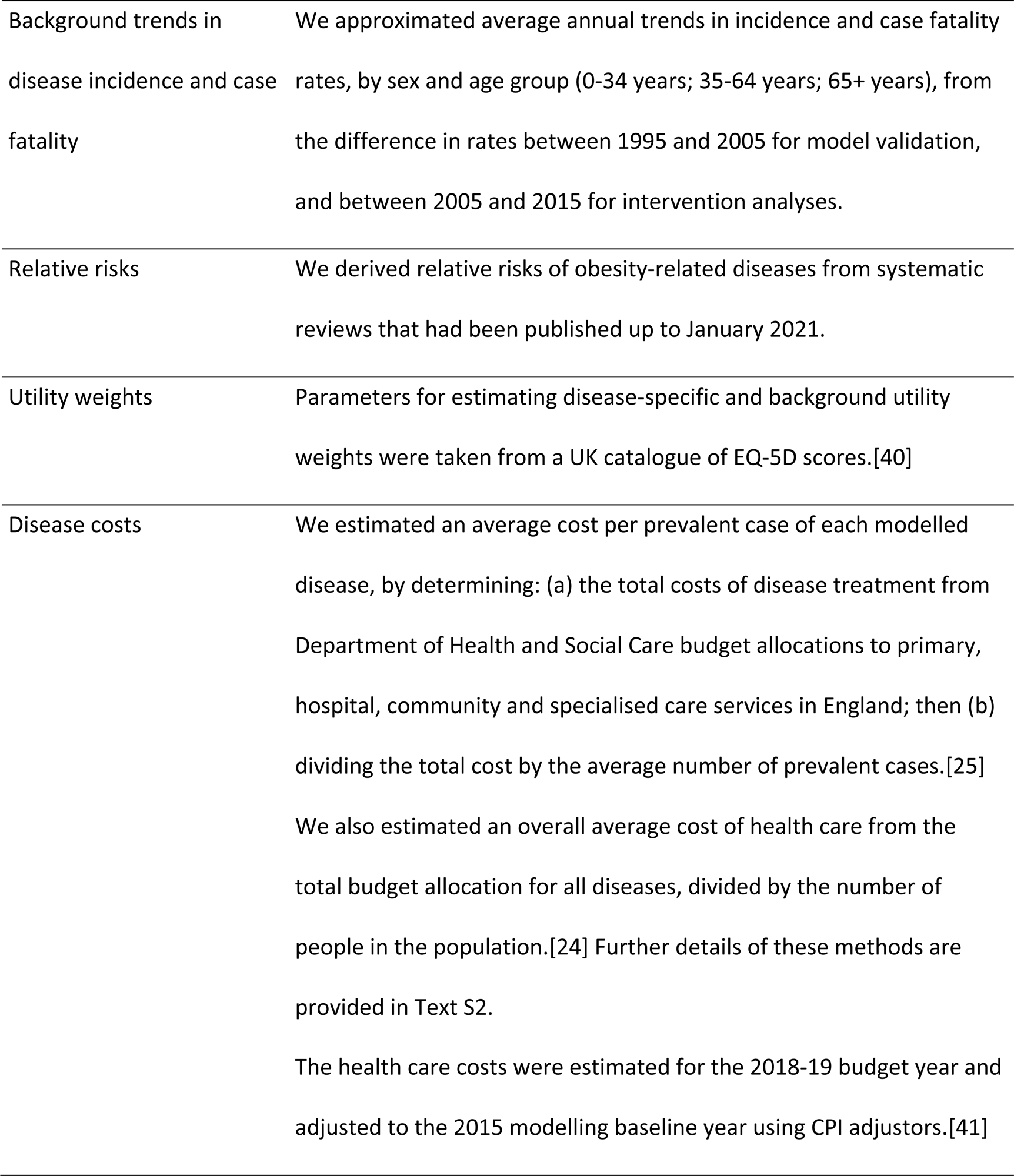
PRIMEtime data inputs.

| PRIMETIME data input | Source |
| --- | --- |
| Population numbers | UK population numbers were determined by 5-year age group and sex from the Human Mortality Database.[24] Future cohorts of 0-4 year olds were derived by sex from the Office for National Statistics (principal projection).[38] |
| All-cause mortality rates | UK all-cause mortality rates were taken from the Human Mortality Database,[24] by single year of age and sex. |
| Background trends in overweight and obesity | Predicted trends in overweight and obesity were taken from models fitted to 27 years of Health Survey for England data, by Cobiac et al.[8] We assumed BMI trends in PRIMETIME would follow the non-linear model in base case analyses and the linear model in sensitivity analyses. |
| Disease rates – incidence, case fatality and starting prevalence | Incidence and case fatality rates were derived from Global Burden of Disease[39] estimates of incidence, prevalence and mortality, using *disbayes*.[27] Spline models were used to smooth data by age. |
| Background trends in disease incidence and case fatality | We approximated average annual trends in incidence and case fatality rates, by sex and age group (0-34 years; 35-64 years; 65+ years), from the difference in rates between 1995 and 2005 for model validation, and between 2005 and 2015 for intervention analyses. |
| Relative risks | We derived relative risks of obesity-related diseases from systematic reviews that had been published up to January 2021. |
| Utility weights | Parameters for estimating disease-specific and background utility weights were taken from a UK catalogue of EQ-5D scores.[40] |
| Disease costs | We estimated an average cost per prevalent case of each modelled disease, by determining: (a) the total costs of disease treatment from Department of Health and Social Care budget allocations to primary, hospital, community and specialised care services in England; then (b) dividing the total cost by the average number of prevalent cases.[25]<br>We also estimated an overall average cost of health care from the total budget allocation for all diseases, divided by the number of people in the population.[24] Further details of these methods are provided in Text S2.<br>The health care costs were estimated for the 2018-19 budget year and adjusted to the 2015 modelling baseline year using CPI adjustors.[41] |

#### Model validation

To validate the epidemiological modelling, we additionally populated PRIMEtime with UK data for a baseline year of 2005, then ran the model forward in time to 2015 and compared simulated prevalence estimates for all cardio-metabolic diseases and cancers with other published data. For comparison we used: (1) GBD estimates of prevalence in the year 2015;^7^ and (2) estimates of prevalence for the period 2010-2015, from an analysis of linked clinical datasets by Kuan et al.[42]

#### Intervention evaluation

To simulate impact of the interventions on obesity prevalence, we ran an open cohort analysis from 2015 to 2050, adding new entrants to the population based on UK population projections. To simulate overall impact on population health (QALYs) and health care costs, we ran a closed cohort analysis over the lifetime of the population alive in 2015. QALYs and costs were discounted using HM Treasury rates of 3.5% costs and 1.5% health (0 to 30 years), 3% costs and 1.29% health (31-75 years) and 2.5% costs and 1.07% health (76-125 years).[43] We also calculated the net monetary benefit of each intervention and the combination of interventions, which reflects the value of each option in monetary terms, assuming a willingness-to-pay threshold for health.[44] For these calculations we assumed a value of £60,000 per QALY, as recommended by HM Treasury.[45]

## Results

### Model validation

Figure 2 illustrates the results of the model validation runs, and full results can be found in Table S1 (PRIMEtime vs. GBD) and Table S2 (PRIMEtime vs. Kuan et al) in the supplementary appendix (Text S3). Overall, estimates from PRIMEtime were slightly higher than estimates from GBD for 13 out of 14 diseases, and slightly lower than estimates from Kuan et al for 12 out of 13 diseases (Kuan et al did not have a comparable disease category for hypertensive heart disease). The absolute difference in the prevalent proportion of the population with disease was less than 1 percentage point for 12 out of 14 diseases in comparison with the GBD estimates, and for 11 out of the 13 diseases in the comparison with the Kuan et al estimates. We found the largest percentage differences for diabetes (Kuan et al 2.7 percentage points lower; GBD 2.5 percentage points higher) and atrial fibrillation and flutter (Kuan et al 2.5 percentage points higher). The mean absolute percentage error across all outcomes was 0.12 percentage points.

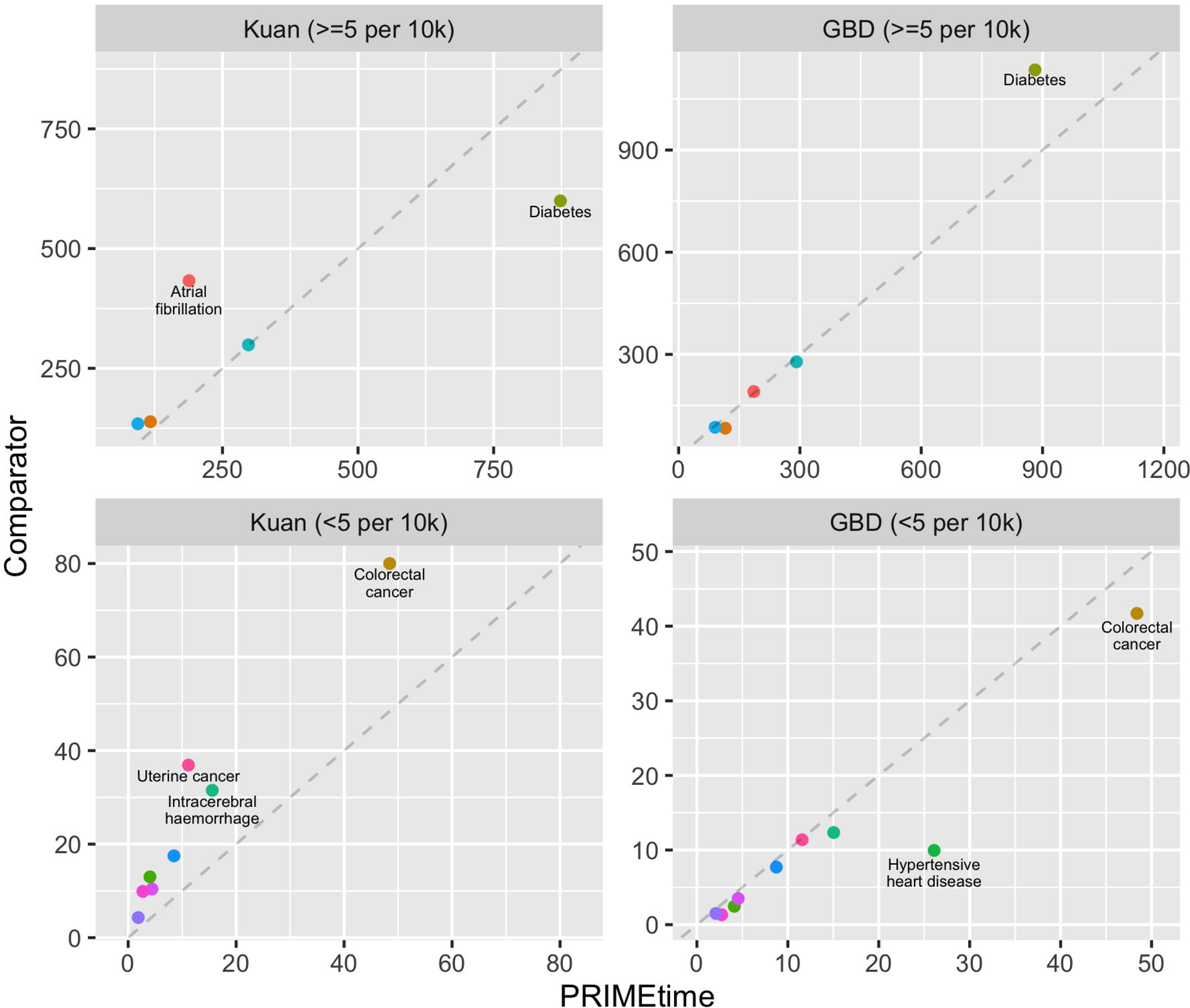

### Prevalence of overweight and obesity

The modelling shows that all of the interventions are likely to reduce population prevalence of overweight and obesity over time, but the pattern of impact varies between interventions (Figure 3). Restricting television advertising of unhealthy foods has the largest impact in reducing overweight and obesity overall, with effects concentrated during childhood. The reduction in prevalence starts at around 0.31 percentage points (95% uncertainty interval: 0.021 to 0.58) for males and 0.34 percentage points (0.023 to 0.65) for females, then increases over time as more children are born and exposed to the intervention. The sugar-sweetened beverage tax also has a slightly smaller impact; benefiting both children and adults, it achieves a reduction in prevalence of overweight and obesity of around 0.19 percentage points (0.058 to 0.31) for males and 0.20 percentage points (0.063 to 0.34) for females initially, and remains relatively stable through time (Text S3 – Table S3). The weight loss program, while very effective for the obese adult population who participate in the program, has the smallest overall effect on population prevalence of overweight and obesity, initially reducing prevalence by just 0.022 percentage points (0.015 to 0.030) for males and 0.027 percentage points (0.018 to 0.037) for females, with lessening benefit over time due to weight regain among participants. The best outcomes are achieved by combining the interventions, which leads to an initial reduction in population prevalence of overweight and obesity of 0.51 percentage point (0.20 to 0.82) for males and 0.57 percentage points (0.22 to 0.90) for females. This reduction increases over time, to around 0.81 percentage points (0.21 to 1.4) for males and 0.95 percentage points (0.24 to 1.7) for females by the end of the simulation in 2050.

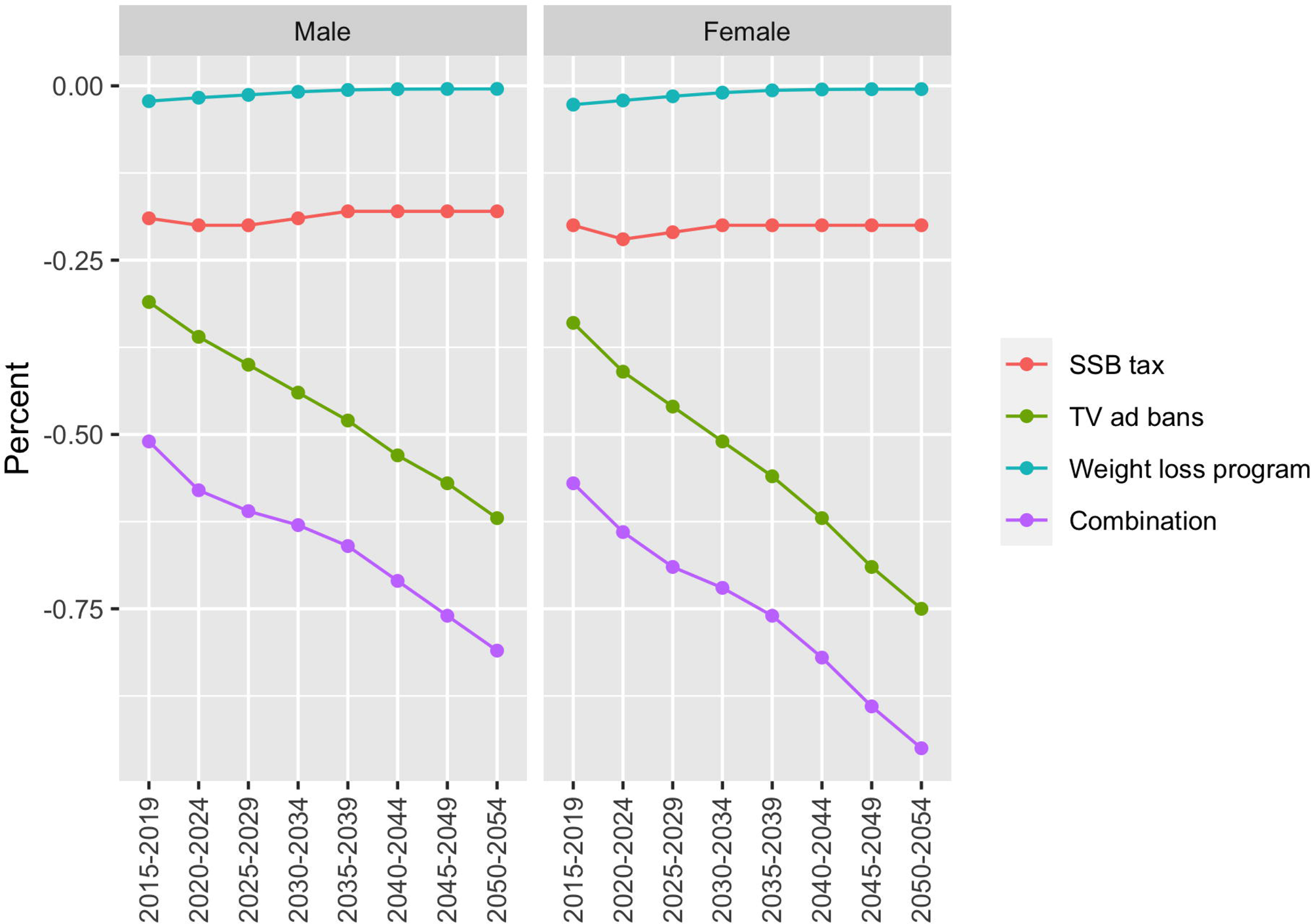

### Cases of disease

Due to the modelled improvements in overweight and obesity prevalence, all of the modelled interventions lead to a reduction in incidence of disease (Table 3). The size of the impact of each intervention, in the first ten years of implementation is strongly influenced by the nature of the target population. The sugar-sweetened beverage tax, which is designed to have an effect across all groups in the population, has the largest benefits in reducing disease in the first ten years. The biggest impact is on cases of type 2 diabetes, followed by cardiovascular diseases and cancers. While restricting television advertising of unhealthy foods to children has larger impacts on overweight and obesity than the SSB tax, it has relatively little effect on cases of disease in the first ten years, since it is targeting children, who will not experience the benefits of disease prevention until some years into the future. The weight loss program, on the other hand, has immediate benefits in reducing cases of type 2 diabetes, cardiovascular diseases and cancers, since it is targeting obese adults, who are at highest risk of developing obesity-related diseases. When combined, the interventions are predicted to prevent 15,000 (95% uncertainty interval: 6,600 to 24,000) cases of type 2 diabetes, 4,900 (2,200 to 7,700) cases of cardiovascular diseases and 460 (220 to 720) cases of cancer in the first ten years of implementation.

**Table 3.**
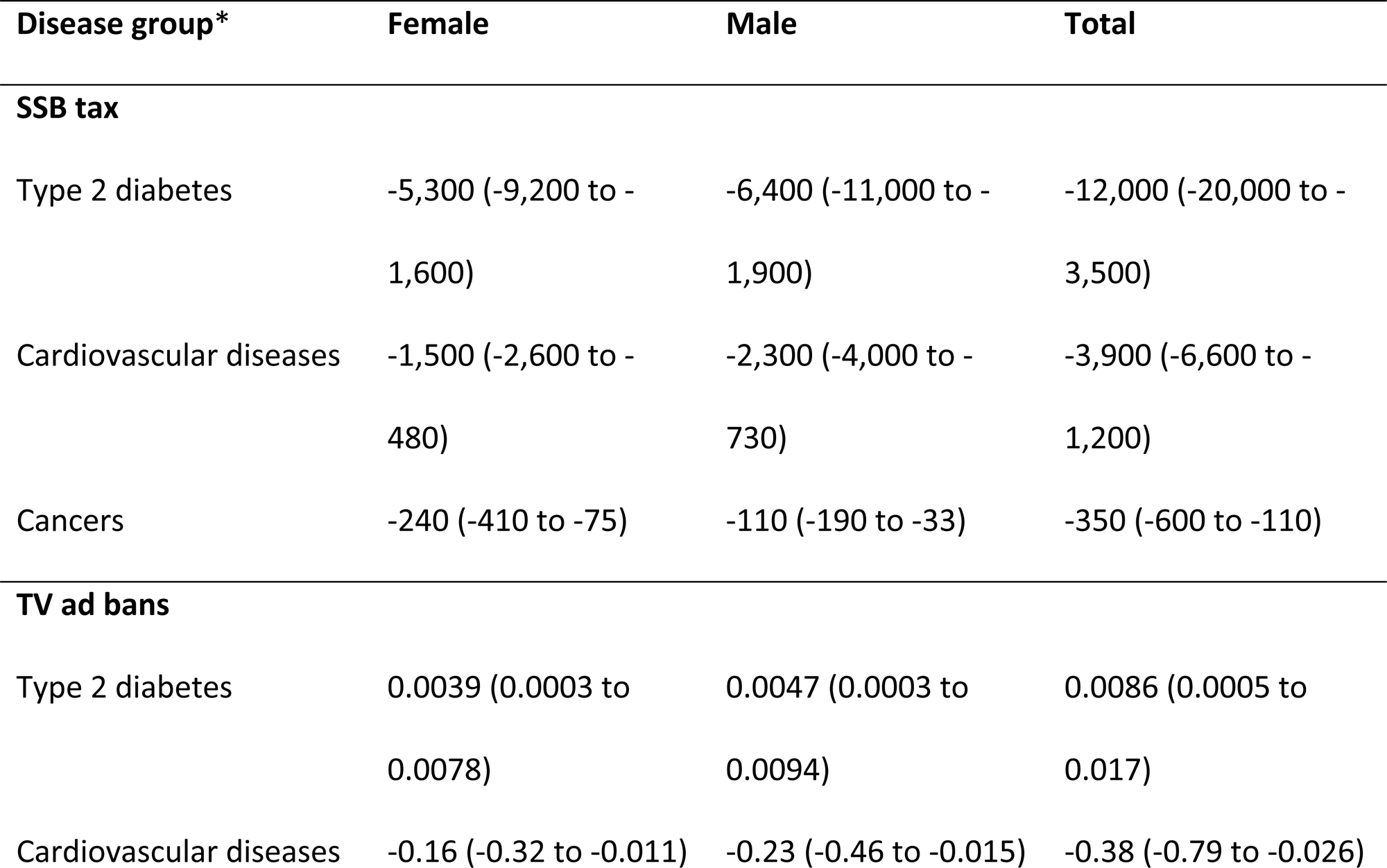
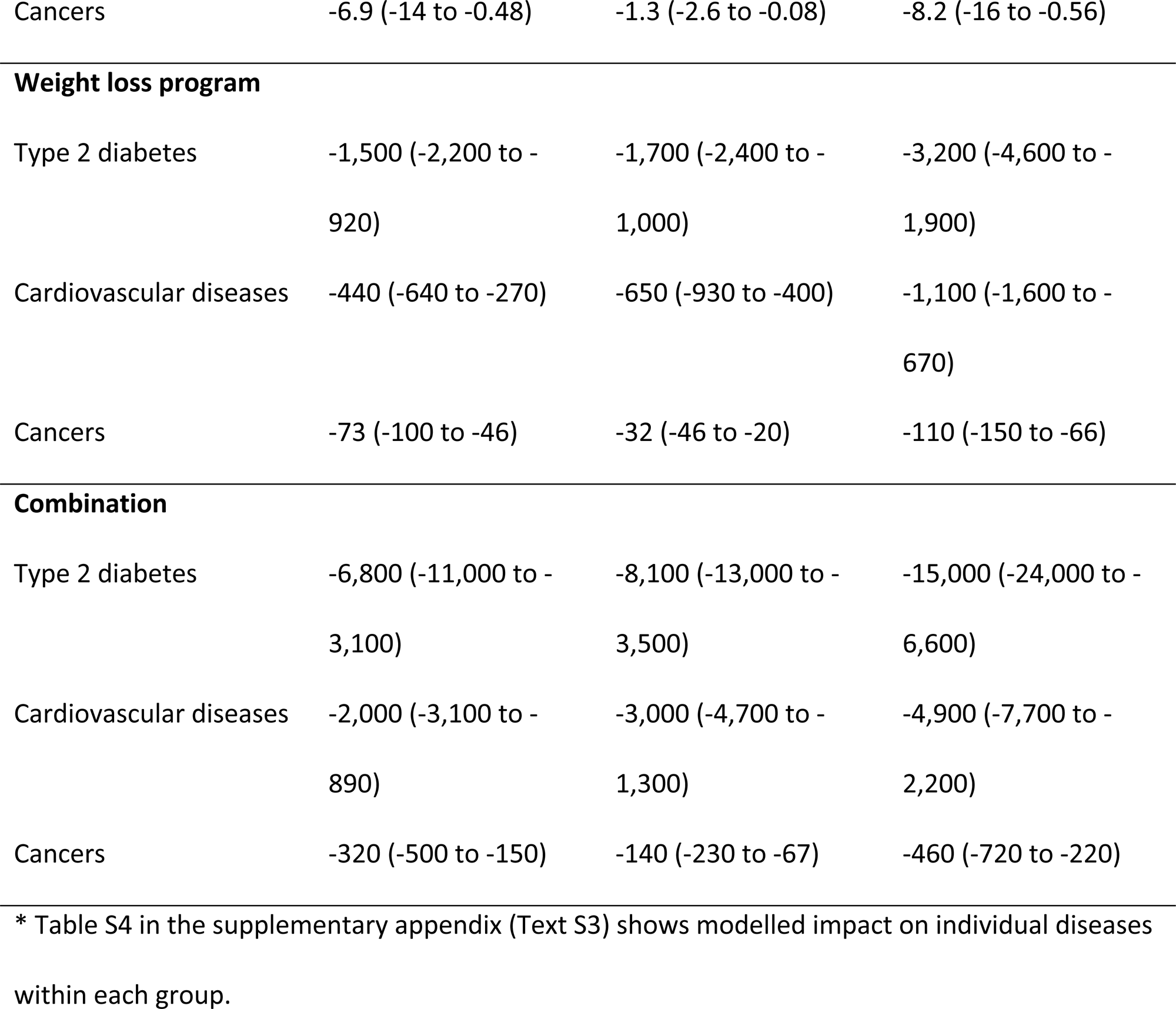
Impact of the intervention scenarios on incident cases of disease in the first ten years (rounded to two significant figures).

### Population health and health care costs

When modelled over the lifetime of the UK population, all of the interventions lead to a health gain for the population and a reduction in health care spending, and all interventions have a positive net monetary benefit from a health sector perspective (Table 4). The combination of interventions is estimated to produce a population health gain of 519,000 (201,000 to 837,000) QALYs and offset £299 (£121 to £515) million in health sector spending, achieving a net monetary benefit of £31,400 (12,200 to 50,700) million.

**Table 4.** Lifetime impact of the intervention scenarios on population health and health care costs, and the net monetary benefit of the scenarios.

| Scenario | Health impact (QALYs) | Cost offsets (£million) | Net monetary benefit (£million) |
| --- | --- | --- | --- |
| SSB tax | 200,000 (61,200 to 345,000) | -128 (-237 to -37.5) | 12,100 (3,710 to 20,900) |
| TV ad bans | 302,000 (19,900 to 588,000) | -159 (-331 to -11.2) | 18,300 (1,200 to 35,600) |
| Weight loss program | 18,600 (8,580 to 33,300) | -12.2 (-26.2 to -2.82) | 1,130 (519 to 2,020) |
| Combination | 519,000 (201,000 to 837,000) | -299 (-515 to -121) | 31,400 (12,200 to 50,700) |

### Uncertainty

Uncertainty around intervention effects on sugar intake (SSB tax), calorie intake (TV ad bans) or weight change (Weight loss program) have the biggest influence on uncertainty in the model predictions of health gain, health sector cost offsets and net monetary benefit (Figure 4). The uncertainty in relative risks of obesity-related diseases, utility values quantifying quality of life, and disease unit costs are less influential. Results are similar when the interventions are evaluated individually (Text S3 – Figure S1).

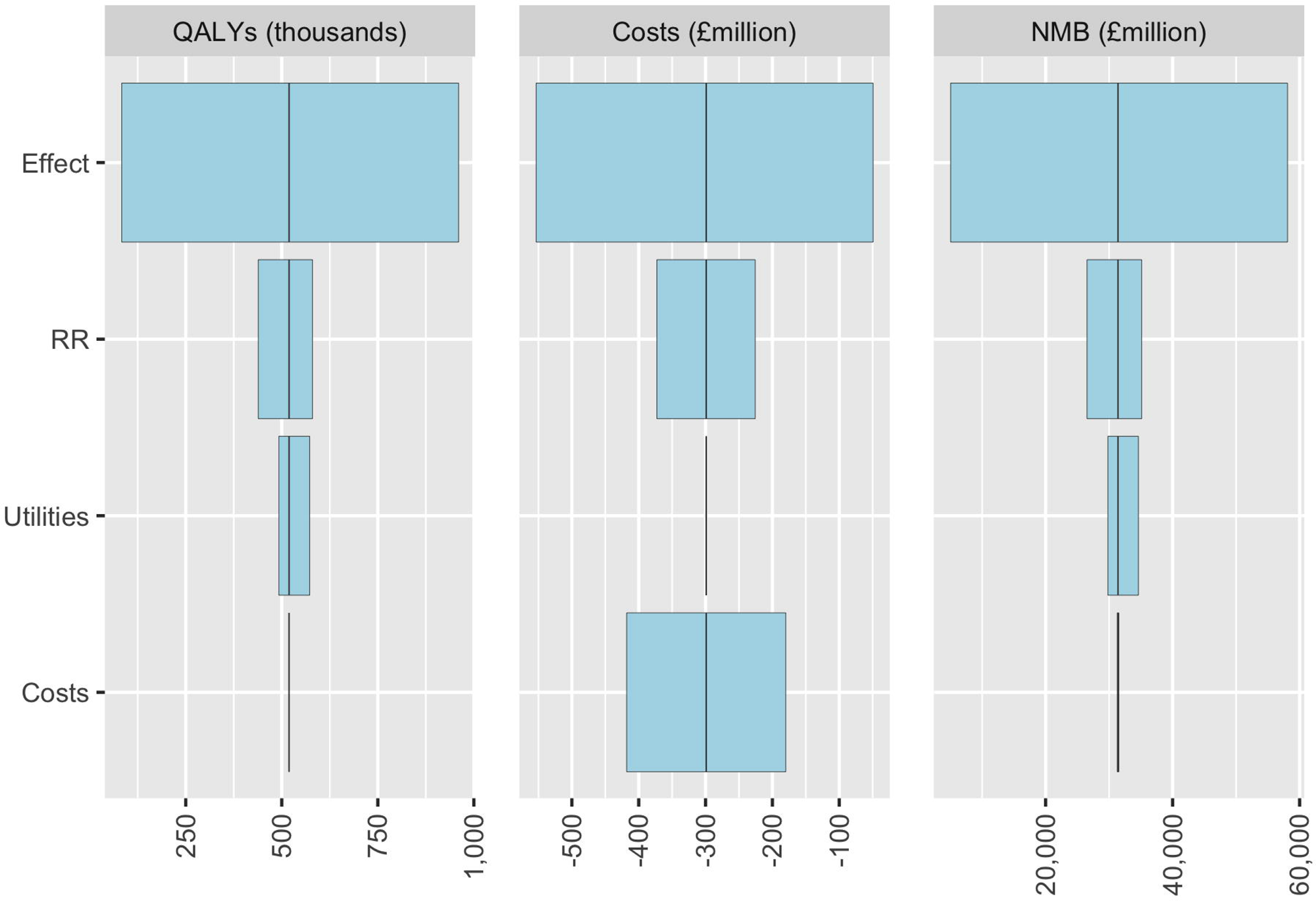
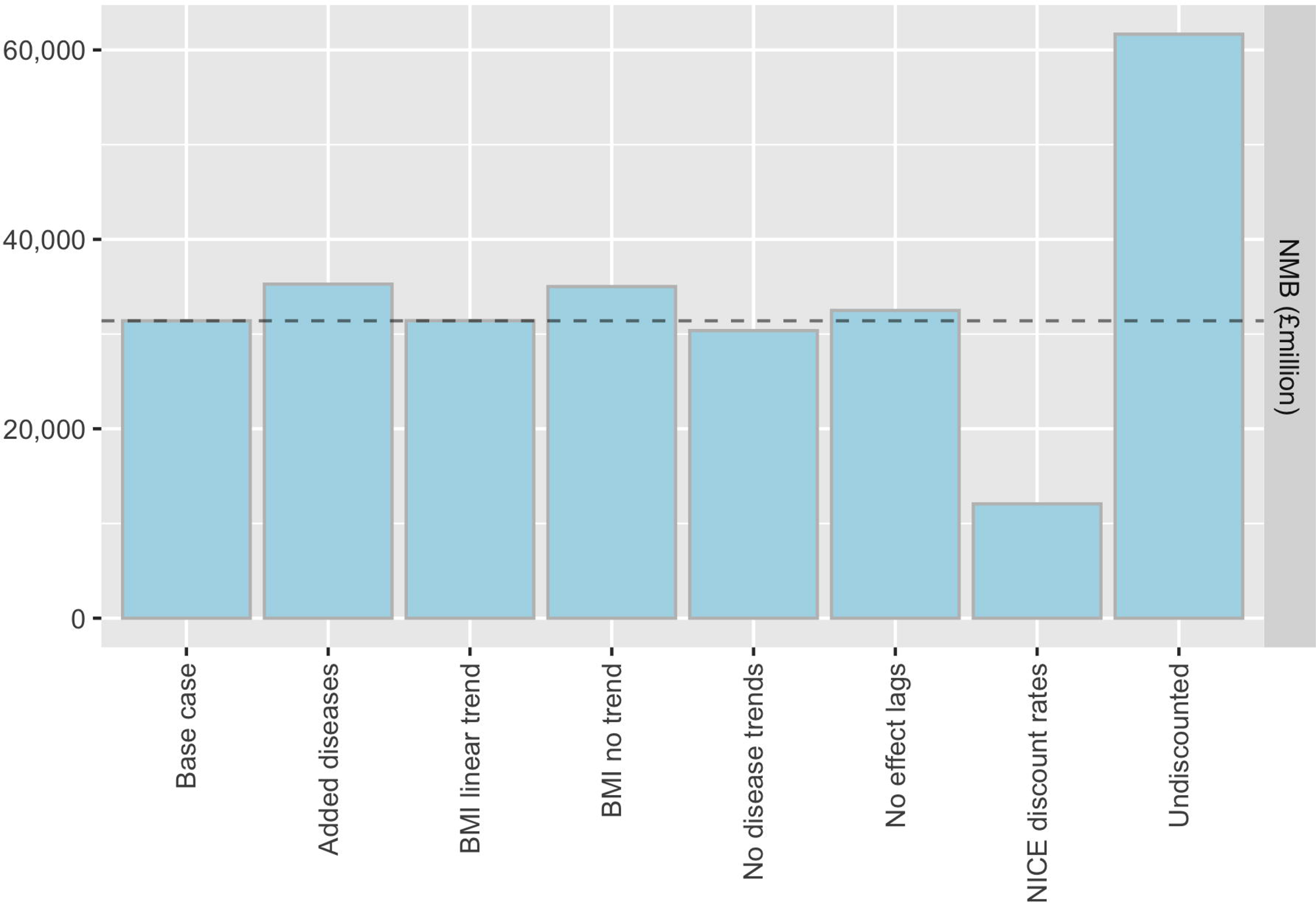

### Sensitivity

The modelled net monetary benefit remained positive in all sensitivity scenarios we evaluated (Text S3 – Figure S2). The magnitude of net monetary benefit is most strongly influenced by the preferred discount rates. The outcome is more favourable with lower or no discounting, since this effectively gives more weight to future health gains and averted health care costs. Whereas the net monetary benefit is less favourable (although still positive) with a higher discount rate overall, or when the rate of discounting of health gains is increased more than costs, (e.g. when changing to NICE rates). Net monetary benefit does not vary substantially with variations in the modelled background trends in BMI distribution, background trends in disease incidence and case fatality rates, and assumed lags in effect of BMI changes on disease incidence. Increasing the number of modelled obesity-related diseases included in the simulation does influence the magnitude of impact on health and health care costs, but the overall impact on net monetary benefit is small.

## Discussion

### Main findings and interpretation

In this paper we have described the PRIMEtime model, a tool for evaluating health and economic impacts of diet and obesity policies. The model uses routinely available epidemiological data to simulate the effects of changes in diet and obesity on 19 non-communicable diseases, over the lifetime of the population, or out to the year 2050 if including future birth cohorts. From these simulations, the model can estimate impact of a policy on population health (overweight and obesity prevalence, cases of disease averted, quality-adjusted life years gained), health and social care costs, and economic measures (net monetary benefit, cost-effectiveness ratios).

The PRIMEtime modelling of a SSB tax, a ban on television advertising of unhealthy foods, and a weight loss program illustrates the potentially wide variation in impact of interventions on prevalence of overweight and obesity, and subsequent changes in health and the need for health care. This variation is due to both the characteristics of the intervention and population that is targeted.

While reductions in TV advertising of unhealthy foods to children has the largest impact in reducing prevalence of overweight and obesity, the SSB tax has much larger and more immediate benefits in reducing disease. The weight loss program also has immediate benefits in improving health despite relatively small benefits in reducing overweight and obesity on a population-scale. The variability in impact, both by age and over time, demonstrates the importance of using models to determine how best to combine intervention strategies to achieve maximum population benefit.

From a health sector perspective, the modelling shows that all three policies will likely have a positive net monetary benefit. This would suggest that all three of the obesity interventions are a good investment for HM Treasury. However, even when combined these three interventions are predicted to reduce prevalence of overweight and obesity by less than 1 percentage point by 2050. With almost a third of children and two-thirds of adults currently overweight or obese,[5] much more will be needed to address the problem.

### Study strengths and limitations

While PRIMEtime is not primarily designed for forecasting disease rates, the accuracy of the underlying epidemiological model will nevertheless impact on the absolute magnitude of its outputs, such as QALYs and health care costs. Therefore it is reassuring that in validation PRIMEtime simulated future prevalence of diseases close to estimates from GBD and Kuan et al.[42] (<1 percentage point difference for 85% of estimates). For the majority of diseases, prevalence estimates from PRIMEtime were slightly higher than those from GBD and slightly lower than those from Kuan et al. This may partly relate to differences in the way that the two comparison studies have arrived at their estimates. The GBD collates data from a variety of sources, such as surveys, registries and published studies, and uses statistical models to estimate country-specific disease rates that are regionally and globally consistent. Whereas, Kuan et al have derived prevalence by analysing electronic health records from the primary care Clinical Practice Research Datalink linked with hospital admissions records from Hospital Episode Statistics, using a newly developed system of algorithmic phenotyping to identify cases of disease.[46] The linked databases are considered representative of the English population,[47] which constitutes around 85% of the UK population that we simulated in PRIMEtime.

Other reasons for differences between PRIMEtime predictions and later estimates may include changes in incidence or case fatality rates that were not captured by trend estimates (e.g. due to increased or decreased funding of prevention or treatment practices) and assumptions embedded in the epidemiological modelling. PRIMEtime simulates the epidemiology using a *proportional* multi-state lifetable design.[23] In a standard multi-state lifetable model, an individual or population cohort transition from one state to another (e.g. from healthy, to having disease *X*, to dying from disease *X* or other causes). But as additional diseases are added to the simulation, the model quickly becomes intractable due the need for additional disease states (e.g. having disease *X* only, having disease *Y* only and having diseases *X* and *Y*) and data required to quantify rates of transition between these many states. A *proportional* multi-state lifetable model overcomes this problem by allowing the individual or population cohort to proportionally exist in multiple disease states. This approach potentially has an advantage in modelling health impacts of *preventive* interventions, where a change in population exposure to a risk factor, can influence many diseases. However, implicit in allowing a population cohort to proportionally exist across multiple disease states is the assumption that the diseases are independent. In some cases, this assumption is clearly violated. For example, prevalence of diabetes increases risk of both heart disease[48] and stroke,[49] even when adjusted for baseline cardiovascular risks such as obesity. In PRIMEtime, we adjust for the dependence between diabetes and cardiovascular disease outcomes, by treating diabetes as both a risk factor and a disease in the model structure. But data to explicitly incorporate other possible relationships, such as between cancer prevalence and cardiovascular outcomes, are limited.

While the proportional multi-state lifetable approach does involve some trade-offs, the methods are widely used in modelling tools designed to evaluate health and economic impacts of diet and obesity interventions (e.g.[50–55]). Models that do not take this approach, typically only simulate one disease (e.g. the Coronary heart disease (CHD) policy model[56, 57]) or estimate changes in incidence and/or mortality of diseases without addressing co-morbidity (e.g. the Foresight model,[58]) thus sacrificing capacity to simulate changing effects on quality of life and health care costs through time.

PRIMEtime is primarily designed for scenario evaluation. It is a tool that can be used to compare future impacts of policies, under a common set of assumptions about future unknowns, and to explore uncertainties in potential outcomes, in order to help set priorities for action and better understand risks associated with decisions. Uncertainty bounds reflect best estimates of the likely range of outcomes, but will never capture impacts of future unpredictable events, such as the emergence of new pandemic diseases. Thus, the true strength of simulation is in comparison of alternative policy options under the same set of assumptions. And these assumptions must be updated as knowledge develops. For example, the selection of diet- and obesity-related diseases in PRIMEtime, and quantification of the dose-response relationships, as captured in Table S4 and Table S5 (supplementary appendix Text S1) should be regularly revisited and updated as new systematic reviews and meta-analyses are published.

### Potential policy and research implications

The three interventions we have evaluated in PRIMEtime are all components of the UK government’s latest obesity strategy.[17] The SSB tax was implemented as a Soft Drinks Industry Levy in 2018;[59] and a 9pm watershed on advertising of high fat, salt and sugar foods, is scheduled to come into effect at the end of 2022.[60] While weight loss programs for those who are already obese are recommended, as of mid-2021, there do not appear to be specific plans for added funding. While the modelling indicates that all three are a good investment, the relatively small overall impact on the high prevalence of overweight and obesity in the UK suggests that much more will be required to improve health and reduce the associated costs of care.

A broader range of interventions targeting dietary intake and obesity may be beneficial for health in the UK, but it is likely that we also need to give greater consideration to the structural and systemic changes that are needed in our society to address the underlying drivers of the obesity epidemic. With globalisation has come enormous shifts in the way that we work, travel, produce and distribute food, which have impacted on both our dietary intake and energy expenditure.[61] While the underlying drivers and feedback mechanisms are sometime complex,[62] the last decade has seen big advances in the way that we can collect and make use of data to better understand these forces, as well as big advances in the tools available to build and run models. Since the early 2000s there has been rapid growth in the development and use of models to find cost-effective interventions that directly target physical activity, dietary intake and obesity.[63, 64] But models can potentially now play a much broader role in simulating impacts of policies or trends that impact on the food system or active environment, either directly or indirectly. In the future, integration of health models, such as PRIMEtime, with models the simulate the complexities of our economies and changing climate, may help us better understand and address the underlying drivers of the obesity epidemic.

## Data Availability

All data used to support the PRIMEtime model and the analyses conducted in this paper are available in the public domain, sometimes with licensed agreements from relevant data archives (e.g. Health Survey for England data).

## References

1. Williams G, Fruhbeck G. Obesity: science to practice: John Wiley & Sons; 2009.

2. Caballero B. The global epidemic of obesity: an overview. Epidemiol Rev. 2007;29(1):1–5.

3. The Global Health Observatory: Indicators: World Health Organization; [13 May 2021]. Available from: https://www.who.int/data/gho/data/indicators.

4. Dai H, Alsalhe TA, Chalghaf N, Riccò M, Bragazzi NL, Wu J. The global burden of disease attributable to high body mass index in 195 countries and territories, 1990–2017: An analysis of the Global Burden of Disease Study. PLoS Med. 2020;17(7):e1003198.

5. Health Survey for England. UK: NHS Digital.

6. Theis DR, White M. Is obesity policy in England fit for purpose? Analysis of government strategies and policies, 1992–2020. The Milbank Quarterly. 2021;99(1):126–70.

7. Jebb S, Aveyard P, Hawkes C. The evolution of policy and actions to tackle obesity in England. Obes Rev. 2013;14:42–59.

8. Cobiac LJ, Scarborough P. Modelling future trajectories of obesity and body mass index in England. PLoS ONE. 2021;16(6):e0252072. doi: 10.1371/journal.pone.0252072.

9. Metcalf CJE, Edmunds W, Lessler J. Six challenges in modelling for public health policy. Epidemics. 2015;10:93–6.

10. Webber L, Mytton OT, Briggs ADM, Woodcock J, Scarborough P, McPherson K, et al. The Brighton declaration: the value of non-communicable disease modelling in population health sciences. Eur J Epidemiol. 2014;29(12):867–70. doi: 10.1007/s10654-014-9978-0.

11. Cobiac LJ, Scarborough P, Kaur A, Rayner M. The Eatwell guide: Modelling the health implications of incorporating new sugar and fibre guidelines. PLoS ONE. 2016;11(12):e0167859.

12. Briggs AD, Cobiac LJ, Wolstenholme J, Scarborough P. PRIMEtime CE: a multistate life table model for estimating the cost-effectiveness of interventions affecting diet and physical activity. BMC Health Serv Res. 2019;19(1):485.

13. Kent S, Aveyard P, Astbury N, Mihaylova B, Jebb SA. Is Doctor Referral to a Low-Energy Total Diet Replacement Program Cost-Effective for the Routine Treatment of Obesity? Obesity. 2019;27(3):391–8.

14. Mytton OT, Boyland E, Adams J, Collins B, O’Connell M, Russell SJ, et al. The potential health impact of restricting less-healthy food and beverage advertising on UK television between 05.30 and 21.00 hours: A modelling study. PLoS Med. 2020;17(10):e1003212.

15. Alonso S, Tan M, Wang C, Kent S, Cobiac L, MacGregor GA, et al. Impact of the 2003 to 2018 Population Salt Intake Reduction Program in England: A Modeling Study. Hypertension. 2021;77(4):1086–94.

16. Cobiac LJ, Scarborough P. Modelling the health co-benefits of sustainable diets in the UK, France, Finland, Italy and Sweden. Eur J Clin Nutr. 2019;73(4):624–33.

17. Department of Health and Social Care. Tackling obesity: empowering adults and children to live healthier lives. UK: Department of Health and Social Care, 2020.

18. Morgenstern H, Bursic ES. A method for using epidemiologic data to estimate the potential impact of an intervention on the health status of a target population. J Community Health. 1982;7(4):292–309.

19. Hill AB. The environment and disease: association or causation?: Sage Publications; 1965.

20. Barendregt J, Oortmarssen GJ, Vos T, Murray CJL. A generic model for the assessment of disease epidemiology: the computational basis of DisMod II. Popul Health Metr. 2003;1. doi: 10.1186/1478-7954-1-4.

21. Lawes C, Vander Hoorn S, Law M, Elliott P. High blood pressure. In: Ezzati M, Lopez A, Rodgers A, Murray C, editors. Comparative Quantification of Health Risks: Global and Regional Burden of Disease Attributable to Selected Major Risk Factors. Geneva: World Health Organisation; 2004.

22. Renehan AG, Soerjomataram I, Tyson M, Egger M, Zwahlen M, Coebergh JW, et al. Incident cancer burden attributable to excess body mass index in 30 European countries. Int J Cancer. 2010;126(3):692–702.

23. Barendregt JJ, Van Oortmarssen GJ, Van Hout BA, Van Den Bosch JM, Bonneux L. Coping with multiple morbidity in a life table. Math Popul Stud. 1998;7(1):29–49.

24. University of California Berkeley (USA), Max Planck Institute for Demographic Research (Germany). Human Mortality Database [5 May 2021]. Available from: www.mortality.org.

25. Institute for Health Metrics and Evaluation. Global Health Data Exchange: GBD Results Tool [5 May 2021]. Available from: http://ghdx.healthdata.org/gbd-results-tool.

26. Institute for Health Metrics and Evaluation. Terms and Conditions [5 May 2021]. Available from: http://www.healthdata.org/about/terms-and-conditions.

27. Jackson C. disbayes [5 May 2021]. Available from: https://github.com/chjackson/disbayes.

28. Global Dietary Database: Tufts University; [13 May 2021]. Available from: https://www.globaldietarydatabase.org/.

29. Briggs AD, Scarborough P, Wolstenholme J. Estimating comparable English healthcare costs for multiple diseases and unrelated future costs for use in health and public health economic modelling. PLoS ONE. 2018;13(5):e0197257.

30. Rogers N, Pell D, Mytton O, Penney T, Briggs A, Cummins S, et al. Changes in soft drinks purchased by British households associated with the UK soft drinks industry levy: a controlled interrupted time series analysis. BMJ Open (Under review).

31. Number of households by household size and age of household reference person (HRP), English regions and UK constituent countries, 2019 UK: Office for National Statistics; 2019 [22 June 2019]. Available from: https://www.ons.gov.uk/peoplepopulationandcommunity/birthsdeathsandmarriages/families/adhocs/11520numberofhouseholdsbyhouseholdsizeandageofhouseholdreferencepersonhrpenglishregionsandukconstituentcountries2019.

32. FAO. Food energy - methods of analysis and conversion factors. Rome: Food and Agriculture Organization of the United Nations, 2003.

33. Hartmann-Boyce J, Theodoulou A, Oke JL, Butler AR, Scarborough P, Bastounis A, et al. Association between characteristics of behavioural weight loss programmes and weight change after programme end: systematic review and meta-analysis. bmj. 2021;374.

34. Jebb SA, Astbury NM, Tearne S, Nickless A, Aveyard P. Doctor Referral of Overweight People to a Low-Energy Treatment (DROPLET) in primary care using total diet replacement products: a protocol for a randomised controlled trial. BMJ open. 2017;7(8).

35. Astbury NM, Aveyard P, Nickless A, Hood K, Corfield K, Lowe R, et al. Doctor Referral of Overweight People to Low Energy total diet replacement Treatment (DROPLET): pragmatic randomised controlled trial. bmj. 2018;362.

36. Hall KD, Butte NF, Swinburn BA, Chow CC. Dynamics of childhood growth and obesity: development and validation of a quantitative mathematical model. The lancet Diabetes & endocrinology. 2013;1(2):97–105.

37. Hall KD, Sacks G, Chandramohan D, Chow CC, Wang YC, Gortmaker SL, et al. Quantification of the effect of energy imbalance on bodyweight. Lancet. 2011;378(9793):826–37. PubMed PMID: S0140-6736(11)60812-X.

38. Office for National Statistics. 2016-based National Population Projections 2017 [1 September 2020]. Available from: https://www.ons.gov.uk/peoplepopulationandcommunity/populationandmigration/populationprojections/datasets/z3zippedpopulationprojectionsdatafilesengland.

39. Global Burden of Disease Study 2015 (GBD 2015) Data Resources: Institute for Health Metrics and Evalaution, University of Washington; 2016 [cited 2016 12 December 2016]. Available from: http://ghdx.healthdata.org/gbd-2015.

40. Sullivan PW, Slejko JF, Sculpher MJ, Ghushchyan V. Catalogue of EQ-5D scores for the United Kingdom. Med Decis Making. 2011;31(6):800–4.

41. CPI INDEX 06: HEALTH 2015=100 United Kingdom: Office for National Statistics; [14 May 2021]. Available from: https://www.ons.gov.uk/economy/inflationandpriceindices/timeseries/d7bz/mm23.

42. Kuan V, Denaxas S, Gonzalez-Izquierdo A, Direk K, Bhatti O, Husain S, et al. A chronological map of 308 physical and mental health conditions from 4 million individuals in the English National Health Service. The Lancet Digital Health. 2019;1(2):e63–e77.

43. HM Treasury. The Green Book: Central Government Guidance on Appraisaland Evaluation. UK: HM Treasury, 2018.

44. Briggs A, Claxton K, Sculpher M. Decision modelling for health economic evaluation. New York: Oxford University Press; 2006.

45. Glover D, Henderson J. Quantifying health impacts of government policies2010 21 October 2021. Available from: https://assets.publishing.service.gov.uk/government/uploads/system/uploads/attachment_data/file/216003/dh_120108.pdf.

46. Denaxas S, Gonzalez-Izquierdo A, Direk K, Fitzpatrick NK, Fatemifar G, Banerjee A, et al. UK phenomics platform for developing and validating electronic health record phenotypes: CALIBER. J Am Med Inform Assoc. 2019;26(12):1545–59.

47. Herrett E, Gallagher AM, Bhaskaran K, Forbes H, Mathur R, Van Staa T, et al. Data resource profile: clinical practice research datalink (CPRD). Int J Epidemiol. 2015;44(3):827–36.

48. Peters SA, Huxley RR, Woodward M. Diabetes as risk factor for incident coronary heart disease in women compared with men: a systematic review and meta-analysis of 64 cohorts including 858,507 individuals and 28,203 coronary events. Diabetologia. 2014;57(8):1542–51.

49. Peters SA, Huxley RR, Woodward M. Diabetes as a risk factor for stroke in women compared with men: a systematic review and meta-analysis of 64 cohorts, including 775 385 individuals and 12 539 strokes. Lancet. 2014;383(9933):1973-80.

50. Brown V, Ananthapavan J, Veerman L, Sacks G, Lal A, Peeters A, et al. The potential cost-effectiveness and equity impacts of restricting television advertising of unhealthy food and beverages to Australian children. Nutrients. 2018;10(5):622.

51. Cleghorn C, Jones A, Freeman L, Wilson N. Updated Cost-effectiveness Modelling of a Behavioural Weight Loss Intervention Involving a Primary Care Provider. New Zealand: University of Otago, 2020.

52. Cobiac LJ, Vos T, Veerman JL. Cost-effectiveness of Lighten Up to a Healthy Lifestyle and Weight Watchers. Report for Queensland Health. Centre for Burden of Disease and Cost-Effectiveness, The University of Queensland, 2008.

53. Forster M, Veerman JL, Barendregt JJ, Vos T. Cost-effectiveness of diet and exercise interventions to reduce overweight and obesity. Int J Obes. 2011.

54. Jones AC, Veerman JL, Hammond D. The health and economic impact of a tax on sugary drinks in Canada. Waterloo (ON): University of Waterloo, 2017.

55. Bourke EJ, Veerman JL. The potential impact of taxing sugar drinks on health inequality in Indonesia. BMJ global health. 2018;3(6):e000923.

56. Moran A, Zhao D, Gu D, Coxson P, Chen C-S, Cheng J, et al. The future impact of population growth and aging on coronary heart disease in China: projections from the Coronary Heart Disease Policy Model-China. BMC Public Health. 2008;8(1):1–14.

57. Weinstein MC, Coxson PG, Williams LW, Pass TM, Stason WB, Goldman L. Forecasting coronary heart disease incidence, mortality, and cost: the Coronary Heart Disease Policy Model. Am J Public Health. 1987;77(11):1417–26.

58. McPherson K, Marsh T, Brown M. Foresight. Tackling obesities – modelling future trends in obesity and the impact on health. London: Government Office for Science, 2007.

59. HM Revenue & Customs. Soft Drinks Industry Levy: UK Government; 2016 [cited 2021 7 April 2021]. Available from: https://www.gov.uk/government/publications/soft-drinks-industry-levy/soft-drinks-industry-levy.

60. Introducing further advertising restrictions on TV and online for products high in fat, salt and sugar: government response: UK Government; 24 June 2021 [30 August 2021]. Available from: https://www.gov.uk/government/consultations/further-advertising-restrictions-for-products-high-in-fat-salt-and-sugar/outcome/introducing-further-advertising-restrictions-on-tv-and-online-for-products-high-in-fat-salt-and-sugar-government-response.

61. Kawachi I, Wamala SP, ProQuest E, Oxford University P. Globalization and health. Wamala S, Kawachi I, Kawachi I, Wamala S, editors. New York;Oxford;: Oxford University Press; 2007.

62. Tackling Obesities: Future Choices – Project report. UK: Government Office for Science, 2007.

63. Cobiac LJ, Veerman L, Vos T. The role of cost-effectiveness analysis in developing nutrition policy. Annu Rev Nutr. 2013;33:373–93.

64. Schwander B, Hiligsmann M, Nuijten M, Evers S. Systematic review and overview of health economic evaluation models in obesity prevention and therapy. Expert review of pharmacoeconomics & outcomes research. 2016;16(5):561–70.

