## Supplementary material for "PRIMEtime: an epidemiological model for informing diet and obesity policy": Text S1

### Text S1 – PRIMEtime data inputs

#### 1. Population numbers

*Table S1 UK population by age and sex from the Human Mortality Database[1] (all ages in 2015) and population projections from the Office for National Statistics[2] (future 0-4 year olds)*

| Age group (years) | Intervention analyses | | | Model validation | | |
|---|---|---|---|---|---|---|
| | Year0* | Male | Female | Year0* | Male | Female |
| 0-4 | 2050 | 2,088,536 | 1,990,411 | – | – | – |
| 0-4 | 2045 | 2,069,673 | 1,972,502 | – | – | – |
| 0-4 | 2040 | 1,999,663 | 1,905,857 | – | – | – |
| 0-4 | 2035 | 1,953,494 | 1,861,924 | – | – | – |
| 0-4 | 2030 | 1,968,036 | 1,875,882 | 2020 | 2,006,190 | 1,910,568 |
| 0-4 | 2025 | 1,994,735 | 1,901,451 | 2015 | 2,063,050 | 1,963,632 |
| 0-4 | 2020 | 2,006,190 | 1,910,568 | 2010 | 1,960,543 | 1,871,193 |
| 0-4 | 2015 | 2,063,050 | 1,963,632 | 2005 | 1,755,352 | 1,669,413 |
| 5-9 | 2015 | 1,999,632 | 1,907,222 | 2005 | 1,860,676 | 1,771,490 |
| 10-14 | 2015 | 1,809,767 | 1,725,856 | 2005 | 1,979,193 | 1,886,300 |
| 15-19 | 2015 | 1,970,469 | 1,866,308 | 2005 | 1,973,931 | 1,911,380 |
| 20-24 | 2015 | 2,191,121 | 2,113,257 | 2005 | 1,953,411 | 1,936,362 |
| 25-29 | 2015 | 2,213,208 | 2,203,621 | 2005 | 1,879,089 | 1,892,516 |
| 30-34 | 2015 | 2,171,167 | 2,197,785 | 2005 | 2,109,298 | 2,139,208 |
| 35-39 | 2015 | 2,009,195 | 2,027,365 | 2005 | 2,314,153 | 2,352,797 |
| 40-44 | 2015 | 2,149,871 | 2,195,049 | 2005 | 2,256,961 | 2,304,185 |
| 45-49 | 2015 | 2,292,536 | 2,359,142 | 2005 | 1,986,295 | 2,015,247 |
| 50-54 | 2015 | 2,227,216 | 2,284,486 | 2005 | 1,824,379 | 1,859,121 |
| 55-59 | 2015 | 1,924,116 | 1,972,675 | 2005 | 1,925,996 | 1,968,812 |
| 60-64 | 2015 | 1,716,700 | 1,789,959 | 2005 | 1,505,366 | 1,568,625 |
| 65-69 | 2015 | 1,742,943 | 1,845,273 | 2005 | 1,297,881 | 1,402,969 |
| 70-74 | 2015 | 1,273,261 | 1,406,103 | 2005 | 1,075,857 | 1,257,363 |
| 75-79 | 2015 | 985,818 | 1,165,295 | 2005 | 830,104 | 1,104,028 |
| 80-84 | 2015 | 671,986 | 903,785 | 2005 | 561,950 | 927,136 |
| 85-89 | 2015 | 366,304 | 604,483 | 2005 | 217,373 | 472,565 |
| 90-94 | 2015 | 134,860 | 309,232 | 2005 | 78,864 | 240,969 |
| 95-100 | 2015 | 22,305 | 82,158 | 2005 | 13,891 | 67,683 |

* Year0 reflects the year in which future cohorts enter the PRIMEtime model simulation.

#### 2. All-cause mortality rates

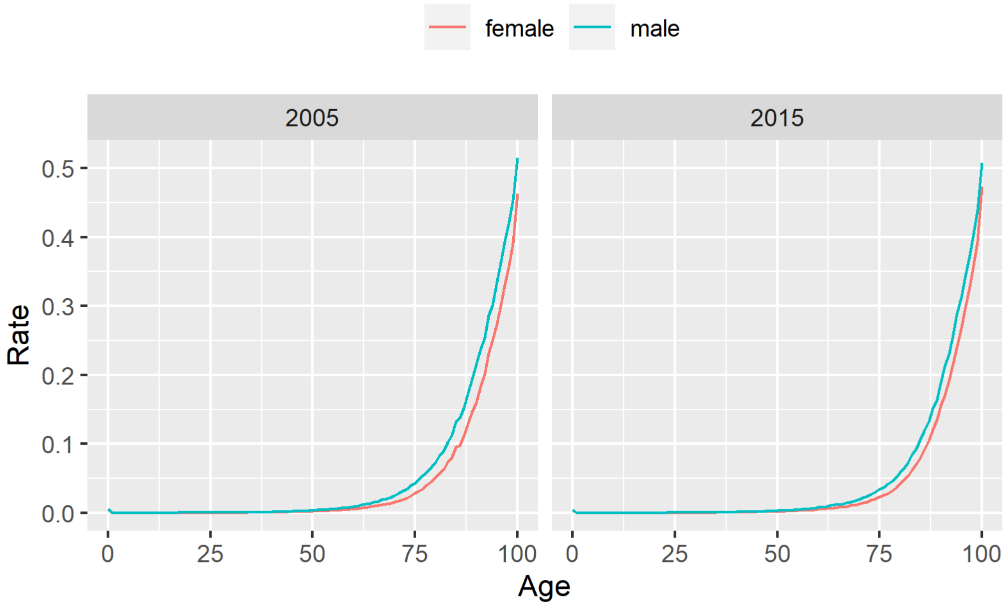

*Figure S1 UK all-cause mortality from the Human Mortality Database[1] for 2005 (model validation) and 2015 (intervention analyses)*

#### 3. Background trends in overweight and obesity

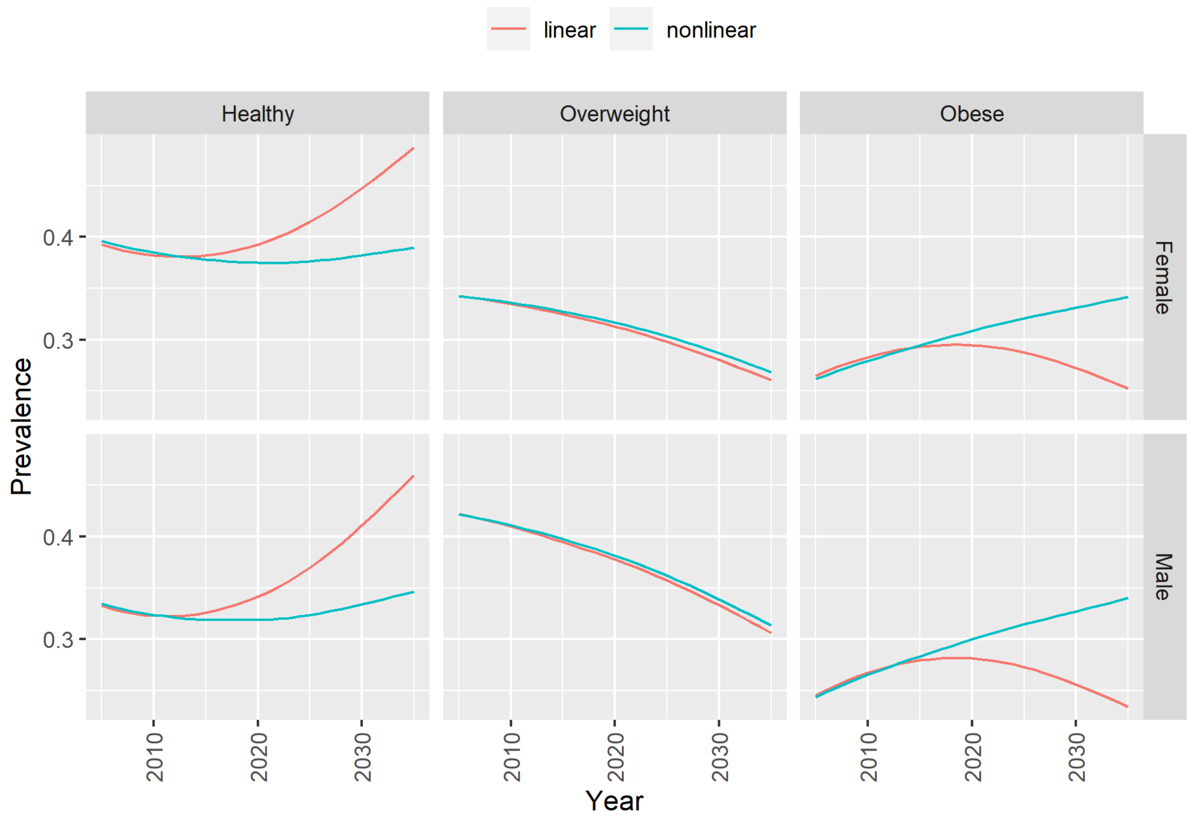

*Figure S2 Predicted trends in overweight and obesity with non-linear (base case) and linear (sensitivity) models from Cobiac et al.[3]*

#### 4. Disease incidence rates

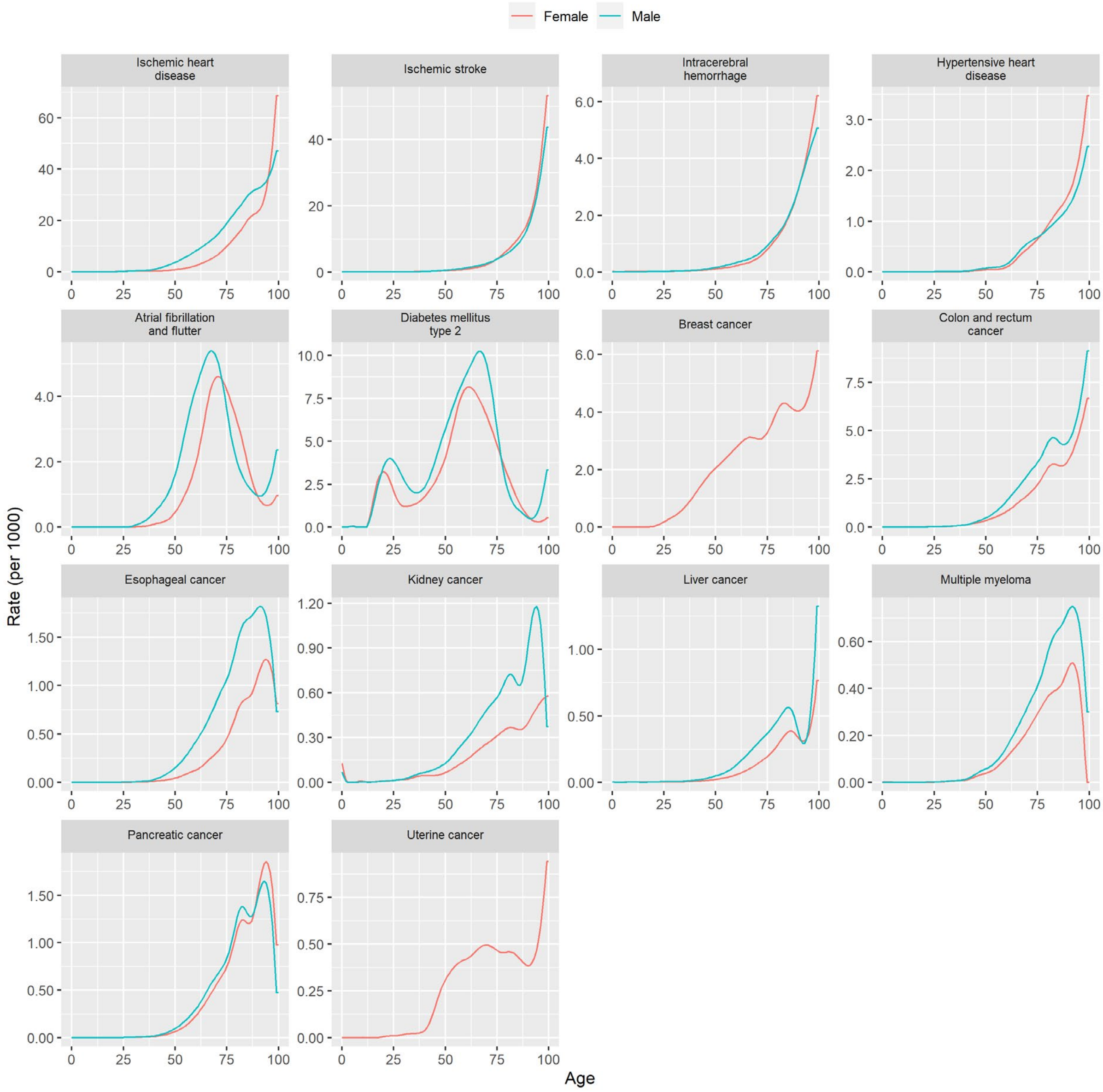

*Figure S3 Incidence rates, derived from Global Burden of Disease[4] estimates using disbayes,[5] – 2005 (Note: graphs are presented on different scales to illustrate each disease in sufficient detail, but this does have the effect of exaggerating variability in rates for some diseases)*

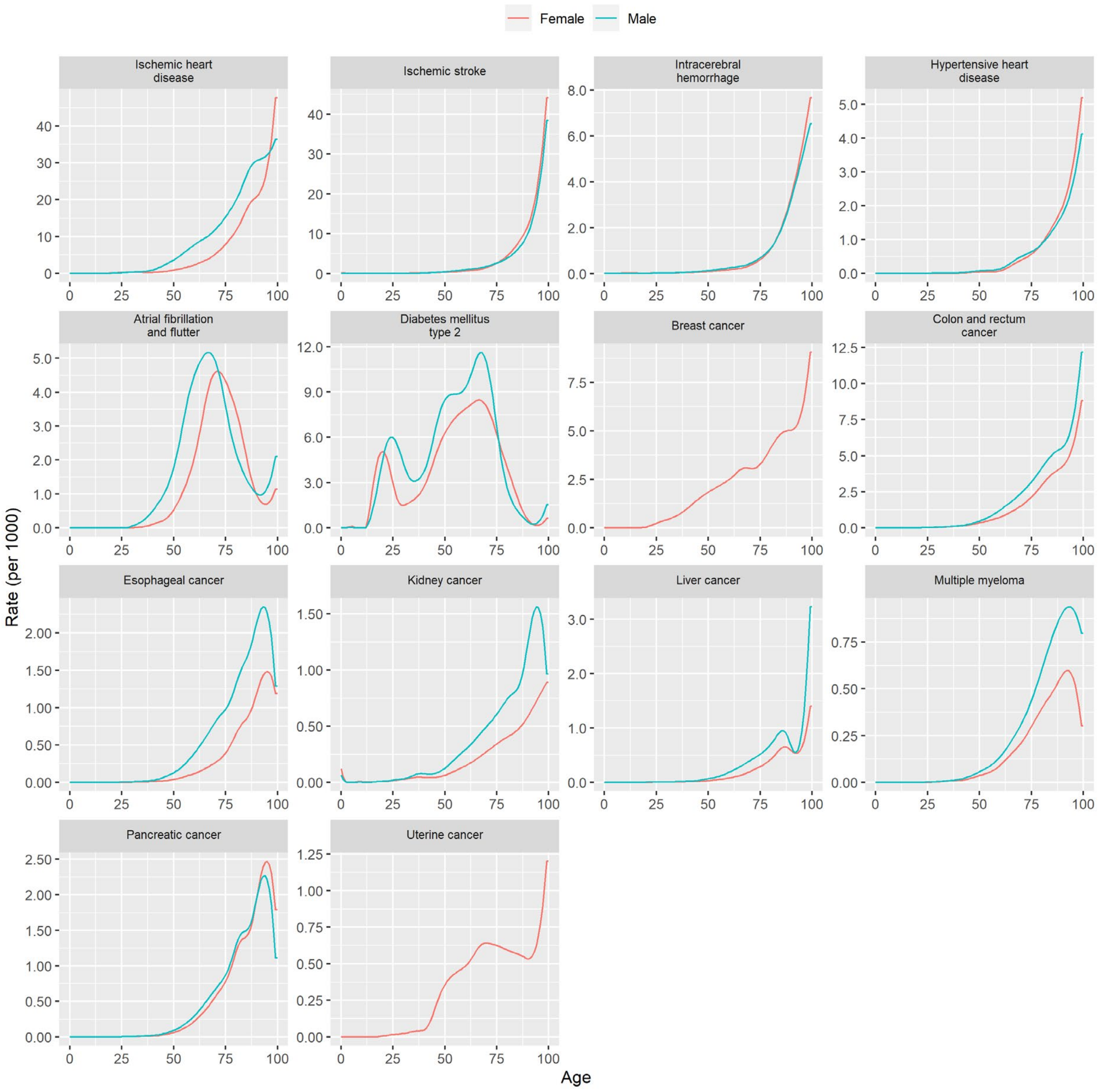

*Figure S4 Incidence rates, derived from Global Burden of Disease[4] estimates using disbayes,[5] – 2015 (Note: graphs are presented on different scales to illustrate each disease in sufficient detail, but this does have the effect of exaggerating variability in rates for some diseases)*

#### 5. Disease case fatality rates

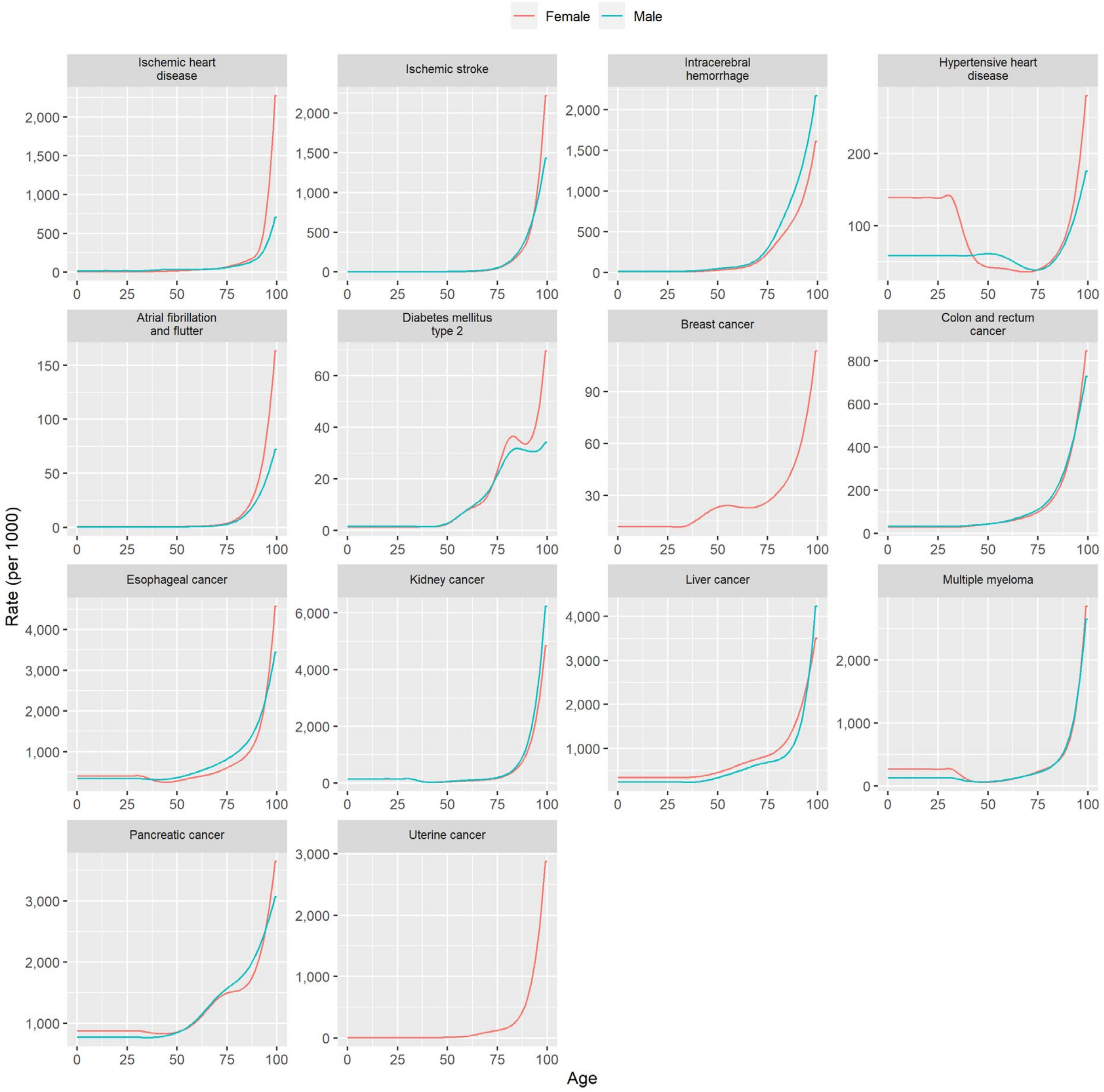

*Figure S5 Case fatality rates, derived from Global Burden of Disease[4] estimates using disbayes,[5] – 2005 (Note: graphs are presented on different scales to illustrate each disease in sufficient detail, but this does have the effect of exaggerating variability in rates for some diseases)*

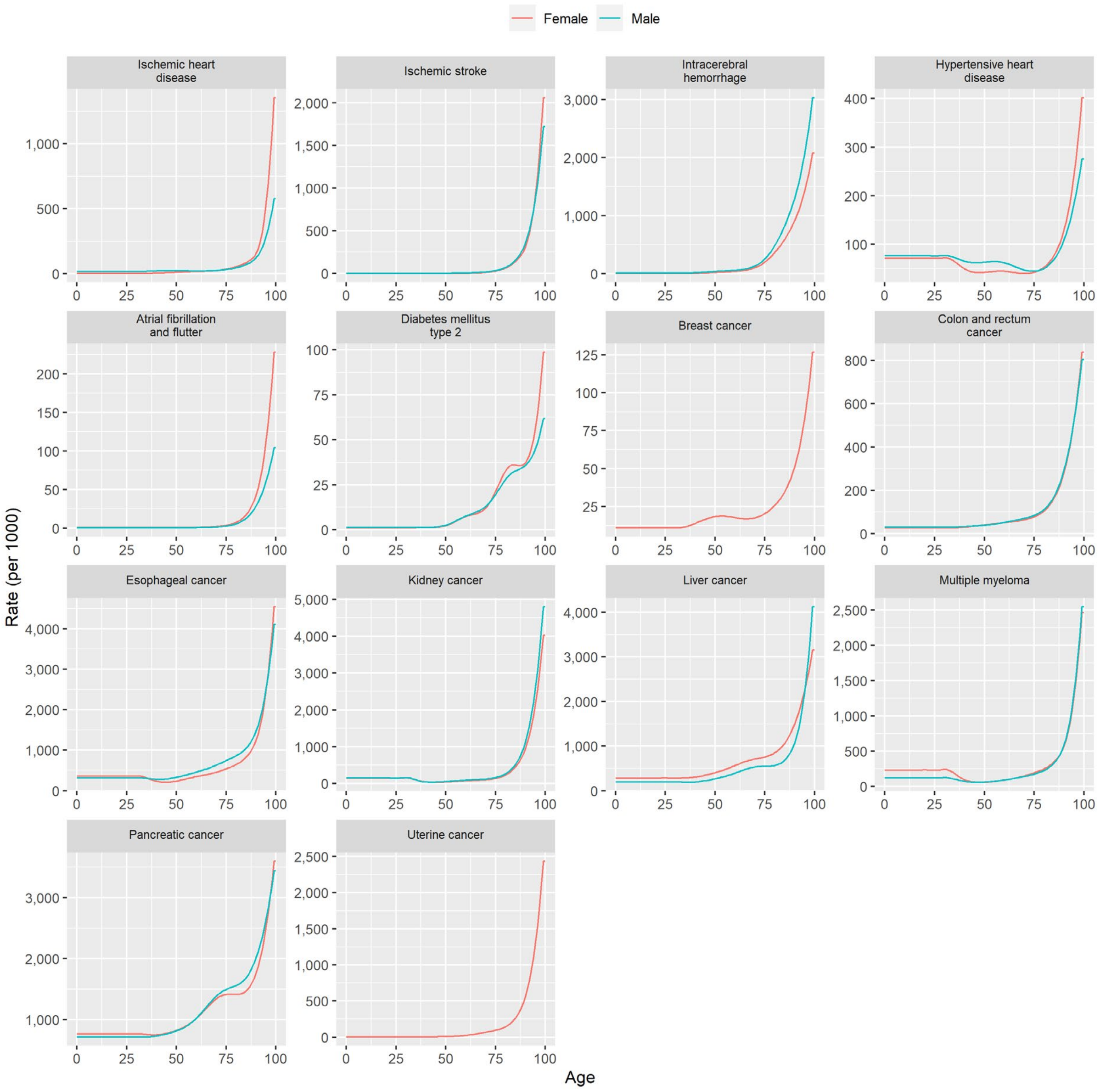

*Figure S6 Case fatality rates, derived from Global Burden of Disease[4] estimates using disbayes,[5] – 2015 (Note: graphs are presented on different scales to illustrate each disease in sufficient detail, but this does have the effect of exaggerating variability in rates for some diseases)*

#### 6. Disease prevalence

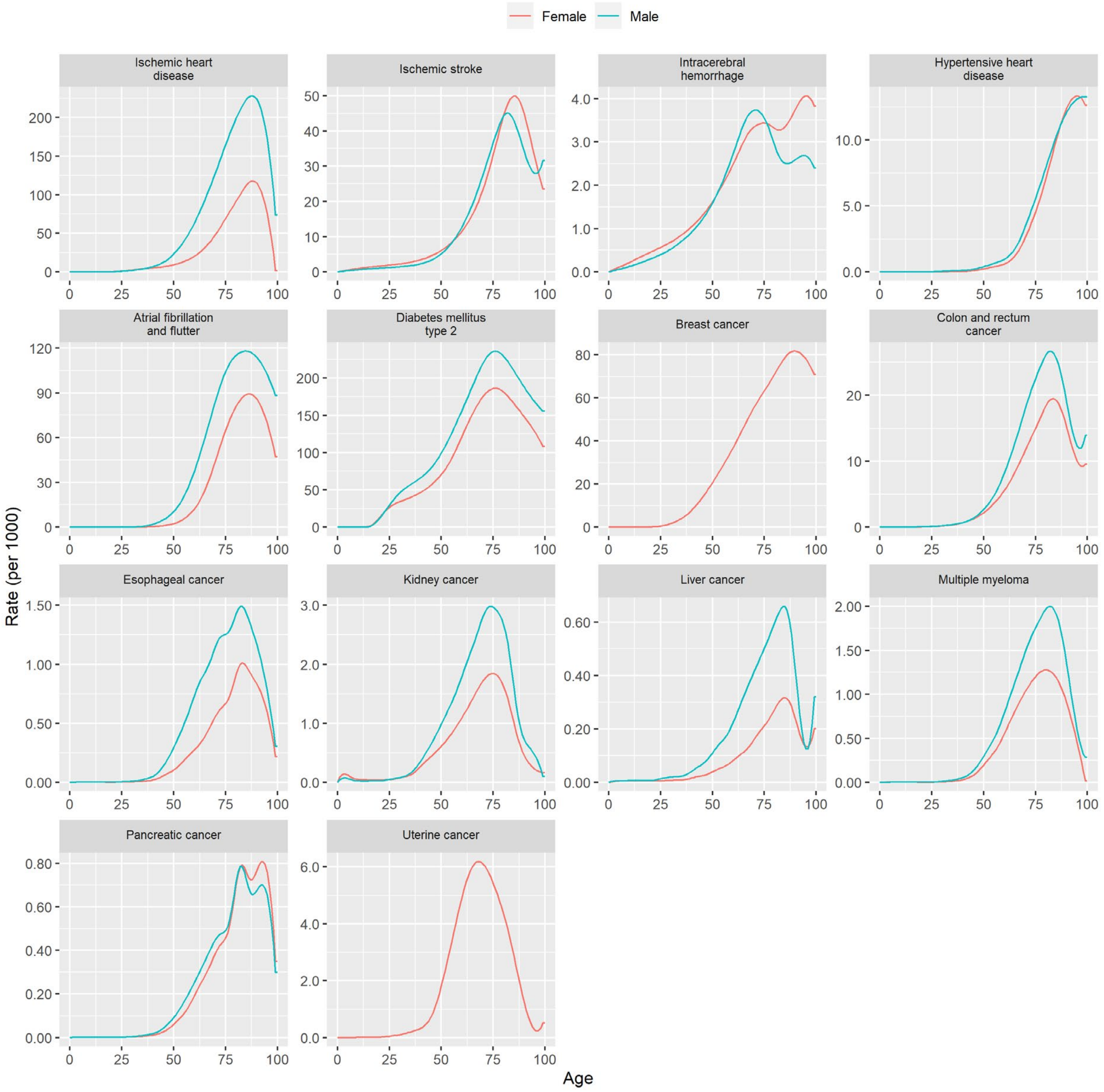

*Figure S7 Starting prevalence rates, derived from Global Burden of Disease[4] estimates using disbayes,[5] – 2005 (Note: graphs are presented on different scales to illustrate each disease in sufficient detail, but this does have the effect of exaggerating variability in rates for some diseases)*

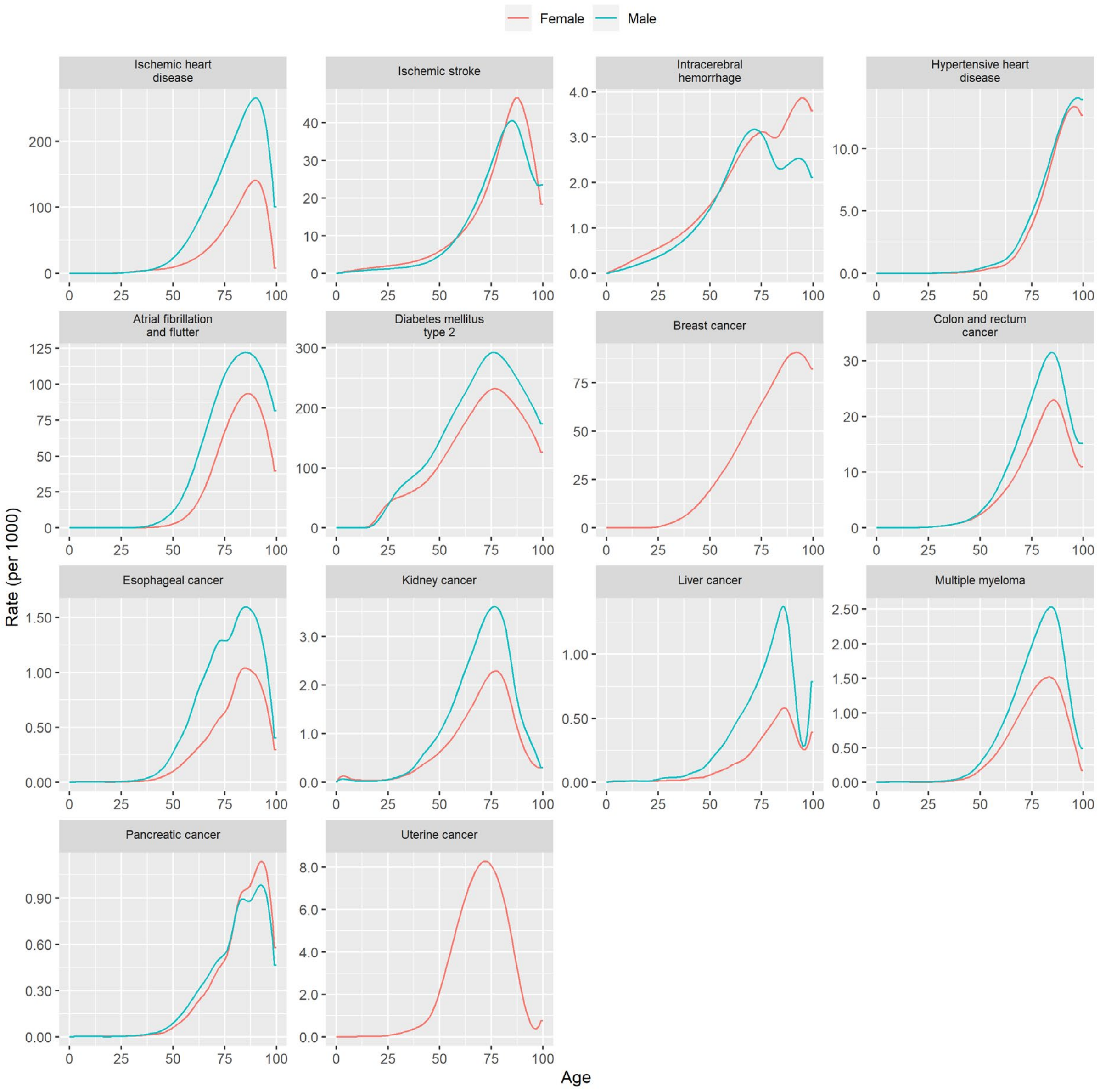

*Figure S8 Starting prevalence rates, derived from Global Burden of Disease[4] estimates using disbayes,[5] – 2015 (Note: graphs are presented on different scales to illustrate each disease in sufficient detail, but this does have the effect of exaggerating variability in rates for some diseases)*

#### 7. Background trends in disease incidence and case fatality

*Table S2 Annual trends in incidence rates, by sex and age group*

| Cause | Female | | | Male | | |
|---|---|---|---|---|---|---|
| | 0-34 | 35-64 | 65+ | 0-34 | 35-64 | 65+ |
| **1995-2005** | | | | | | |
| Ischaemic heart disease | -0.0123 | -0.0256 | -0.0302 | -0.0223 | -0.0223 | -0.0284 |
| Ischaemic stroke | -0.0083 | -0.0279 | -0.0300 | -0.0099 | -0.0248 | -0.0334 |
| Intracerebral haemorrhage | -0.0080 | -0.0328 | -0.0074 | -0.0134 | -0.0231 | -0.0147 |
| Hypertensive heart disease | 0.0514 | 0.0289 | 0.1270 | 0.0548 | 0.0757 | 0.1290 |
| Atrial fibrillation and flutter | -0.0043 | -0.0008 | -0.0050 | 0.0091 | -0.0010 | -0.0072 |
| Diabetes mellitus type 2 | 0.0945 | 0.0022 | 0.0144 | 0.0901 | 0.0022 | 0.0472 |
| Breast cancer | -0.0192 | -0.0065 | 0.0008 | NA | NA | NA |
| Colon and rectum cancer | 0.0063 | -0.0103 | -0.0059 | 0.0073 | -0.0080 | -0.0039 |
| Oesophageal cancer | -0.0026 | -0.0013 | 0.0023 | -0.0014 | 0.0061 | 0.0085 |
| Kidney cancer | 0.0093 | 0.0025 | 0.0218 | 0.0178 | 0.0058 | 0.0140 |
| Liver cancer | 0.0231 | 0.0294 | 0.0419 | 0.0389 | 0.0371 | 0.0428 |
| Multiple myeloma | 0.0123 | 0.0011 | 0.0074 | 0.0130 | 0.0036 | 0.0063 |
| Pancreatic cancer | -0.0027 | -0.0006 | 0.0076 | -0.0025 | -0.0043 | 0.0006 |
| Uterine cancer | 0.0422 | 0.0292 | 0.0303 | NA | NA | NA |
| **2005-2015** | | | | | | |
| Ischaemic heart disease | 0.0052 | 0.0013 | -0.0205 | -0.0030 | 0.0007 | -0.0156 |
| Ischaemic stroke | 0.0033 | -0.0172 | -0.0320 | -0.0007 | -0.0180 | -0.0318 |
| Intracerebral haemorrhage | -0.0051 | -0.0202 | -0.0041 | -0.0091 | -0.0203 | -0.0121 |
| Hypertensive heart disease | 0.0185 | -0.0027 | 0.0034 | 0.0086 | 0.0028 | 0.0037 |
| Atrial fibrillation and flutter | 0.0069 | 0.0142 | 0.0023 | 0.0069 | 0.0120 | -0.0019 |
| Diabetes mellitus type 2 | 0.0379 | 0.0261 | 0.0220 | 0.0376 | 0.0303 | 0.0131 |
| Breast cancer | 0.0112 | -0.0073 | 0.0042 | NA | NA | NA |
| Colon and rectum cancer | 0.0359 | 0.0013 | 0.0008 | 0.0295 | 0.0003 | -0.0021 |
| Oesophageal cancer | 0.0105 | -0.0066 | -0.0078 | 0.0166 | -0.0046 | -0.0028 |
| Kidney cancer | 0.0147 | 0.0015 | 0.0136 | 0.0214 | 0.0057 | 0.0093 |
| Liver cancer | 0.0291 | 0.0372 | 0.0418 | 0.0359 | 0.0372 | 0.0398 |
| Multiple myeloma | 0.0020 | -0.0056 | 0.0056 | 0.0028 | 0.0009 | 0.0079 |
| Pancreatic cancer | 0.0102 | 0.0024 | 0.0076 | 0.0126 | 0.0030 | 0.0068 |
| Uterine cancer | 0.0395 | 0.0209 | 0.0269 | NA | NA | NA |

*Table S3 Annual trends in case fatality rates, by sex and age group*

| Cause | Female | | | Male | | |
|---|---|---|---|---|---|---|
| | 0-34 | 35-64 | 65+ | 0-34 | 35-64 | 65+ |
| **1995-2005** | | | | | | |
| Ischaemic heart disease | -0.0403 | -0.0433 | -0.0498 | -0.0179 | -0.0340 | -0.0491 |
| Ischaemic stroke | -0.0335 | -0.0489 | -0.0085 | -0.0362 | -0.0505 | -0.0248 |
| Intracerebral haemorrhage | -0.0328 | -0.0284 | 0.0013 | -0.0164 | -0.0173 | -0.0020 |
| Hypertensive heart disease | 0.2510 | 0.1150 | -0.1930 | 0.0831 | 0.0732 | -0.0849 |
| Atrial fibrillation and flutter | -0.0072 | -0.0059 | 0.0147 | 0.0285 | 0.0066 | 0.0102 |
| Diabetes mellitus type 2 | -0.0147 | -0.0156 | -0.0138 | -0.0128 | -0.0181 | -0.0183 |
| Breast cancer | -0.0238 | -0.0256 | -0.0141 | NA | NA | NA |
| Colon and rectum cancer | -0.0162 | -0.0207 | -0.0163 | -0.0184 | -0.0228 | -0.0215 |
| Oesophageal cancer | -0.0189 | -0.0151 | -0.0105 | -0.0154 | -0.0112 | -0.0097 |
| Kidney cancer | -0.0030 | -0.0165 | -0.0167 | -0.0016 | -0.0223 | -0.0196 |
| Liver cancer | -0.0238 | -0.0178 | -0.0140 | -0.0243 | -0.0240 | -0.0206 |
| Multiple myeloma | -0.0239 | -0.0265 | -0.0160 | -0.0290 | -0.0296 | -0.0216 |
| Pancreatic cancer | -0.0082 | -0.0058 | -0.0036 | -0.0056 | -0.0037 | -0.0023 |
| Uterine cancer | 0.0269 | -0.0130 | -0.0059 | NA | NA | NA |
| **2005-2015** | | | | | | |
| Ischaemic heart disease | -0.0023 | -0.0348 | -0.0503 | 0.0127 | -0.0325 | -0.0467 |
| Ischaemic stroke | -0.0647 | -0.0424 | -0.0292 | -0.0547 | -0.0320 | -0.0285 |
| Intracerebral haemorrhage | -0.0234 | -0.0178 | -0.0055 | -0.0142 | -0.0137 | -0.0052 |
| Hypertensive heart disease | -0.0668 | -0.0200 | 0.0170 | 0.0257 | 0.0088 | 0.0184 |
| Atrial fibrillation and flutter | 0.0091 | -0.0061 | 0.0098 | -0.0174 | -0.0126 | 0.0081 |
| Diabetes mellitus type 2 | -0.0084 | -0.0017 | -0.0048 | -0.0122 | -0.0013 | -0.0038 |
| Breast cancer | -0.0055 | -0.0198 | -0.0182 | NA | NA | NA |
| Colon and rectum cancer | -0.0112 | -0.0094 | -0.0186 | -0.0108 | -0.0117 | -0.0206 |
| Oesophageal cancer | -0.0117 | -0.0140 | -0.0104 | -0.0058 | -0.0084 | -0.0094 |
| Kidney cancer | -0.0060 | -0.0086 | -0.0175 | -0.0046 | -0.0096 | -0.0134 |
| Liver cancer | -0.0206 | -0.0110 | -0.0099 | -0.0218 | -0.0201 | -0.0195 |
| Multiple myeloma | -0.0132 | -0.0078 | -0.0129 | -0.0041 | -0.0083 | -0.0144 |
| Pancreatic cancer | -0.0133 | -0.0054 | -0.0061 | -0.0068 | -0.0039 | -0.0047 |
| Uterine cancer | -0.0197 | -0.0096 | -0.0163 | NA | NA | NA |

#### 8. Relative risks of disease

*Table S4 Relative risks of modelled obesity-related diseases*

| Disease | Population subgroup | Units/Category | Mean relative risk (95% CI) | Source |
|---|---|---|---|---|
| Ischaemic heart disease | 35-44 | per 5kg/m$^{32}$ | 1.66 (1.51 to 1.84) | Singh et al 2013[6] |
| | 45-54 | per 5kg/m$^{31}$ | 1.55 (1.46 to 1.64) | |
| | 55-64 | per 5kg/m$^{30}$ | 1.44 (1.4 to 1.48) | |
| | 65-74 | per 5kg/m$^{29}$ | 1.35 (1.32 to 1.38) | |
| | 75-84 | per 5kg/m$^{28}$ | 1.26 (1.2 to 1.32) | |
| | 85+ | per 5kg/m$^{27}$ | 1.14 (1.04 to 1.26) | |
| Ischaemic stroke | 35-44 | per 5kg/m$^{26}$ | 1.86 (1.67 to 2.08) | Singh et al 2013[6] |
| | 45-54 | per 5kg/m$^{25}$ | 1.67 (1.53 to 1.81) | |
| | 55-64 | per 5kg/m$^{24}$ | 1.5 (1.4 to 1.6) | |
| | 65-74 | per 5kg/m$^{23}$ | 1.35 (1.28 to 1.41) | |
| | 75-84 | per 5kg/m$^{22}$ | 1.21 (1.16 to 1.26) | |
| | 85+ | per 5kg/m$^{21}$ | 1.04 (0.96 to 1.12) | |
| Intracerebral haemorrhage | 35-44 | per 5kg/m$^{20}$ | 2.54 (1.96 to 3.28) | Singh et al 2013[6] |
| | 45-54 | per 5kg/m$^{19}$ | 2.1 (1.66 to 2.66) | |
| | 55-64 | per 5kg/m$^{18}$ | 1.75 (1.44 to 2.13) | |
| | 65-74 | per 5kg/m$^{17}$ | 1.48 (1.29 to 1.71) | |
| | 75-84 | per 5kg/m$^{16}$ | 1.3 (1.21 to 1.4) | |
| | 85+ | per 5kg/m$^{15}$ | 1.05 (0.92 to 1.2) | |
| Hypertensive heart disease | 35-44 | per 5kg/m$^{14}$ | 2.15 (0.8 to 5.78) | Singh et al 2013[6] |
| | 45-54 | per 5kg/m$^{13}$ | 2.02 (0.97 to 4.21) | |
| | 55-64 | per 5kg/m$^{12}$ | 1.9 (1.17 to 3.07) | |
| | 65-74 | per 5kg/m$^{11}$ | 1.81 (1.45 to 2.26) | |
| | 75-84 | per 5kg/m$^{10}$ | 1.63 (1.53 to 1.74) | |
| | 85+ | per 5kg/m$^{9}$ | 1.45 (1.05 to 2.01) | |
| Diabetes mellitus 2 | 35-44 | per 5kg/m$^{8}$ | 3.07 (2.28 to 4.15) | Singh et al 2013[6] |
| | 45-54 | per 5kg/m$^{7}$ | 2.66 (2.15 to 3.3) | |
| | 55-64 | per 5kg/m$^{6}$ | 2.32 (2.04 to 2.63) | |
| | 65-74 | per 5kg/m$^{5}$ | 2.03 (1.95 to 2.11) | |
| | 75-84 | per 5kg/m$^{4}$ | 1.7 (1.61 to 1.79) | |
| | 85+ | per 5kg/m$^{3}$ | 1.38 (1.23 to 1.56) | |

| | | | | |
|---|---|---|---|---|
| Atrial fibrillation and flutter | – | per 5kg/$m^2$ | 1.28 (1.2 to 1.38) | Aune et al 2017[7] |
| Gallbladder and biliary diseases | – | per 5kg/$m^1$ | 1.63 (1.49 to 1.78) | Aune et al 2015[8] |
| Colon and rectum cancer | – | per 5kg/$m^0$ | 1.05 (1.03 to 1.07) | WCRF 2018[9] |
| Breast cancer | women | per 5kg/$m^1$ | 1.12 (1.09 to 1.15) | WCRF 2018[9] |
| Uterine cancer | women | per 5kg/$m^2$ | 1.54 (1.47 to 1.61) | Kyrgiou et al 2017[10] |
| Oesophageal cancer | adeno-carcinoma | per 5kg/$m^3$ | 1.54 (1.41 to 1.67) | Kyrgiou et al 2017[10] |
| | squamous cell carcinoma | per 5kg/$m^4$ | 0.63 (0.53 to 0.75) | |
| Kidney cancer | men | per 5kg/$m^5$ | 1.24 (1.17 to 1.32) | Kyrgiou et al 2017[10] |
| | women | per 5kg/$m^6$ | 1.33 (1.25 to 1.42) | |
| Pancreatic cancer | – | per 5kg/$m^2$ | 1.1 (1.06 to 1.14) | Kyrgiou et al 2017[10] |
| Multiple myeloma | – | per 5kg/$m^2$ | 1.12 (1.09 to 1.15) | Kyrgiou et al 2017[10] |
| Liver cancer | – | per 5kg/$m^2$ | 1.3 (1.16 to 1.46) | WCRF 2018[9] |
| Asthma* | <18 years | Healthy | 1 | Azizpour et al 2018[11] |
| | | Overweight | 1.64 (1.13 to 2.38) | |
| | | Obese | 1.92 (1.39 to 2.65) | |
| | 18+ years | Healthy | 1 | Beuther et al 2007[12] |
| | | Overweight | 1.38 (1.17 to 1.62) | |
| | | Obese | 1.92 (1.43 to 2.59) | |
| Low back pain* | – | Healthy | 1 | Shiri et al 2010[13] |
| | | Overweight | 1.08 (0.9 to 1.29) | |
| | | Obese | 1.42 (1.11 to 0.181) | |
| Osteoarthritis knee* | 50+ years | Healthy | 1 | Silverwood et al 2015[14] |
| | | Overweight | 1.98 (1.57 to 2.2) | |
| | | Obese | 2.66 (2.15 to 3.28) | |
| Osteoarthritis hip* | – | per 5kg/$m^2$ | 1.11 (1.07 to 1.16) | Jiang et al 2011[15] |
| Depressive disorders* | – | Healthy | 1 | Amiri et al 2018[16] |
| | – | Overweight | 1.04 (0.99 to 1.11) | |
| | – | Obese | 1.15 (1.06 to 1.25) | |

* Disease included in sensitivity analyses only

*Table S5 Relative risk of ischaemic heart disease and ischaemic stroke in people with type 2 diabetes*

| Disease | Population subgroup | Units/Category | Mean relative risk (95% CI) | Source |
|---|---|---|---|---|
| Ischaemic heart disease | men | Diabetes | 1.85 (1.64 to 2.1) | Peters et al 2014[17] |
| | women | Diabetes | 2.63 (2.27 to 3.06) | |
| Ischaemic stroke | men | Diabetes | 1.83 (1.6 to 2.08) | Peters et al 2014[18] |
| | women | Diabetes | 2.28 (1.93 to 2.69) | |

#### 9. Utility weights

*Table S6 Disease-specific utility weights, estimated for the UK by Sullivan et al[19]*

| Disease | ICD9 code | Mean utility (SD) |
|---|---|---|
| Ischaemic heart disease incidence | icd410 | -0.063 (0.013) |
| Ischaemic heart disease prevalence | icd412 | -0.037 (0.026) |
| Ischaemic stroke incidence | icd436 | -0.117 (0.012) |
| Ischaemic stroke prevalence | icd438 | -0.073 (0.024) |
| Intracerebral haemorrhage incidence | icd436 | -0.117 (0.012) |
| Intracerebral haemorrhage prevalence | icd438 | -0.073 (0.024) |
| Hypertensive heart disease | icd401 | -0.046 (0.004) |
| Diabetes mellitus type 2 | icd250 | -0.071 (0.005) |
| Atrial fibrillation and flutter | icd427 | -0.038 (0.007) |
| Colon and rectum cancer | icd153 | -0.067 (0.017) |
| Breast cancer | icd174 | -0.019 (0.014) |
| Uterine cancer | icd202 | -0.010 (0.026) |
| Oesophageal cancer | icd202 | -0.010 (0.026) |
| Kidney cancer | icd189 | -0.048 (0.041) |
| Pancreatic cancer | icd202 | -0.010 (0.026) |
| Multiple myeloma | icd195 | -0.086 (0.027) |
| Liver cancer | icd155 | -0.093 (0.044) |
| Asthma | icd493 | -0.046 (0.006) |
| Low back pain | icd724 | -0.087 (0.006) |
| Osteoarthritis hip | icd715 | -0.114 (0.008) |
| Osteoarthritis knee | icd715 | -0.114 (0.008) |
| Depressive disorders | icd296 | -0.127 (0.010) |
| Gallbladder and biliary diseases | icd574-76* | -0.062 (0.018) |

* weighted by Hospital Episode Statistics admissions primary diagnosis for ICD10 K80-83.[20]

*Table S7 Parameters for estimating background utility weight, , estimated for the UK by Sullivan et al[19]*

| Parameter | Mean utility (SD) |
|---|---|
| Age (continuous years) | -0.00027 (0.00017) |
| Male | 0.0010 (0.00063) |
| Age 10-19 | 0.913 (0.0045) |
| Age 20-29 | 0.905 (0.0021) |
| Age 30-39 | 0.879 (0.0021) |
| Age 40-49 | 0.837 (0.0028) |
| Age 50-59 | 0.798 (0.0035) |
| Age 60-69 | 0.774 (0.0039) |
| Age 70-79 | 0.723 (0.0049) |
| Age 80-89 | 0.657 (0.0075) |

#### 10. Disease costs

*Table S8 Costs of treatment in the National Health Service (see Text S2)*

| Condition | Units | Mean cost (SD)* |
|---|---|---|
| Ischaemic heart disease | prevalent case | £606 (£121) |
| Ischaemic stroke | prevalent case | £1,950 (£390) |
| Intracerebral haemorrhage | prevalent case | £2,563 (£513) |
| Hypertensive heart disease | prevalent case | £103 (£21) |
| Diabetes mellitus type 2 | prevalent case | £187 (£37) |
| Atrial fibrillation and flutter | prevalent case | £195 (£39) |
| Colon and rectum cancer | incident case | £9,204 (£1,841) |
| Breast cancer | incident case | £12,433 (£2,487) |
| Uterine cancer | incident case | £2,060 (£412) |
| Oesophageal cancer | incident case | £2,421 (£484) |
| Kidney cancer | incident case | £4,979 (£996) |
| Pancreatic cancer | incident case | £2,695 (£539) |
| Multiple myeloma | incident case | £22,915 (£4,583) |
| Liver cancer | incident case | £2,172 (£434) |
| Asthma | incident case | £4,186 (£837) |
| Low back pain | incident case | £425 (£85) |
| Osteoarthritis hip | incident case | £13,951 (£2,790) |
| Osteoarthritis knee | incident case | £1,799 (£360) |
| Depressive disorders | incident case | £410 (£82) |
| Gallbladder and biliary diseases | incident case | £372 (£74) |
| Total non-modelled diseases | person | £1,099 (£220) |

* Standard deviation estimated as 20% of point estimate.
** Decayed missing and filled deciduous (dmft) and permanent (DMFT) teeth.

#### References

1. University of California Berkeley (USA), Max Planck Institute for Demographic Research (Germany). Human Mortality Database [5 May 2021]. Available from: https://www.mortality.org.

2. Office for National Statistics. 2016-based National Population Projections 2017 [1 September 2020]. Available from: https://www.ons.gov.uk/peoplepopulationandcommunity/populationandmigration/populationprojections/datasets/z3zippedpopulationprojectionsdatafilesengland.

3. Cobiac LJ, Scarborough P. Modelling future trajectories of obesity and body mass index in England. PLoS ONE. 2021;16(6):e0252072. doi: 10.1371/journal.pone.0252072.

4. Global Burden of Disease Study 2015 (GBD 2015) Data Resources: Institute for Health Metrics and Evalaution, University of Washington; 2016 [cited 2016 12 December 2016]. Available from: http://ghdx.healthdata.org/gbd-2015.

5. Jackson C. disbayes [5 May 2021]. Available from: https://github.com/chjackson/disbayes.

6. Singh GM, Danaei G, Farzadfar F, Stevens GA, Woodward M, Wormser D, et al. The age-specific quantitative effects of metabolic risk factors on cardiovascular diseases and diabetes: a pooled analysis. PLoS ONE. 2013;8(7):e65174.

7. Aune D, Sen A, Schlesinger S, Norat T, Janszky I, Romundstad P, et al. Body mass index, abdominal fatness, fat mass and the risk of atrial fibrillation: a systematic review and dose–response meta-analysis of prospective studies. Eur J Epidemiol. 2017;32(3):181-92.

8. Aune D, Norat T, Vatten LJ. Body mass index, abdominal fatness and the risk of gallbladder disease. Eur J Epidemiol. 2015;30(9):1009-19.

9. World Cancer Research Fund. Diet, Nutrition, Physical Activity and Cancer: a Global Perspective, Third Expert Report. World Cancer Research Fund; American Institute for Cancer Research, 2018.

10. Kyrgiou M, Kalliala I, Markozannes G, Gunter MJ, Paraskevaidis E, Gabra H, et al. Adiposity and cancer at major anatomical sites: umbrella review of the literature. Bmj. 2017;356.

11. Azizpour Y, Delpisheh A, Montazeri Z, Sayehmiri K, Darabi B. Effect of childhood BMI on asthma: a systematic review and meta-analysis of case-control studies. BMC Pediatr. 2018;18(1):1-13.

12. Beuther DA, Sutherland ER. Overweight, obesity, and incident asthma: a meta-analysis of prospective epidemiologic studies. Am J Respir Crit Care Med. 2007;175(7):661-6.

13. Shiri R, Karppinen J, Leino-Arjas P, Solovieva S, Viikari-Juntura E. The association between obesity and low back pain: a meta-analysis. Am J Epidemiol. 2010;171(2):135-54.

14. Silverwood V, Blagojevic-Bucknall M, Jinks C, Jordan J, Protheroe J, Jordan K. Current evidence on risk factors for knee osteoarthritis in older adults: a systematic review and meta-analysis. Osteoarthritis Cartilage. 2015;23(4):507-15.

15. Jiang L, Rong J, Wang Y, Hu F, Bao C, Li X, et al. The relationship between body mass index and hip osteoarthritis: a systematic review and meta-analysis. Joint Bone Spine. 2011;78(2):150-5.

16.Amiri S, Behnezhad S, Nadinlui KB. Body Mass Index (BMI) and risk of depression in adults: A systematic review and meta-analysis of longitudinal studies. Obesity Medicine. 2018;12:1-12.

17.Peters SA, Huxley RR, Woodward M. Diabetes as risk factor for incident coronary heart disease in women compared with men: a systematic review and meta-analysis of 64 cohorts including 858,507 individuals and 28,203 coronary events. Diabetologia. 2014;57(8):1542-51.

18.Peters SA, Huxley RR, Woodward M. Diabetes as a risk factor for stroke in women compared with men: a systematic review and meta-analysis of 64 cohorts, including 775 385 individuals and 12 539 strokes. Lancet. 2014;383(9933):1973-80.

19.Sullivan PW, Slejko JF, Sculpher MJ, Ghushchyan V. Catalogue of EQ-5D scores for the United Kingdom. Med Decis Making. 2011;31(6):800-4.

20.NHS Digital. Hospital Episode Statistics for England: Admitted Patient Care statistics, 2019-20 2020 [21 April 2021]. Available from: https://digital.nhs.uk/data-and-information/publications/statistical/hospital-admitted-patient-care-activity/2019-20.
