## Supplementary material for "PRIMEtime: an epidemiological model for informing diet and obesity policy": Text S2

### Text S2 – Estimating healthcare costs

#### NHS costs of diet- and obesity-related diseases

We follow the method developed by Briggs et al (2018) that uses the ICD-10 codes to estimate comparable disease specific English healthcare costs. In 2018/19, NHS England received £114bn funding from the Department of Health & Social Care (DHSC 2019). 74% of this budget was allocated to the clinical commissioning groups (CCG), which are responsible for commissioning local NHS health care services in England including mental health services, urgent and emergency care, elective hospital care, and community care. Of the remainder, £11.8bn was allocated to primary care, with the rest distributed across specialised services, public health and NHS England administration costs (DHSC, 2019). Specialised services are nationally commissioned healthcare services which are either particularly expensive or have a small patient population. CCG, specialised services and primary care therefore form the majority of diseases-related NHS England spend. Following the cost estimation approach developed by Briggs et al (2018), these healthcare costs in 2018/19 are assigned to different diseases in the following steps:

*Step 1 Allocate the CCG expenditure.*

The programming budgeting (PB) data from NHS England, reports the CCG expenditure across 56 PB categories which are referenced to ICD-10 codes. The total CCG expenditure is assigned to each modelled disease based on their ICD-10 codes. For asthma, the ICD-10 codes used in PRIMEtime are the same as those found in the PB category 11b. Asthma. The total expenditure from this category (£749m) was therefore used as the amount spent by the CCGs on asthma in 2018/19. For other modelled diseases, there are some differences between their respective ICD-codes and those found in a single PB category. Take osteoarthritis hip as an example, its ICD-10 codes M16-M16.9 are only some of the codes under the PB category 15x. Problems of the Musculo skeletal system. The 2018/19 hospital episode statistics (HES) data from NHS Digital is used to estimate the proportion of PB costs that can be allocated to relevant ICD-10 codes. This dataset provides details of all admissions at NHS hospitals in England, allowing us to calculate ratio of admissions from the ICD-10 codes of each modelled disease to admissions from all ICD-10 codes in the relevant PB category. This ratio is then multiplied by the total CCG expenditure under each PB category to estimate disease specific CCG expenditure. In the case of osteoarthritis hip, the ratio of admission is 0.085 as among 1,057,069 patients admitted from the PB category *15x* in 2018/19, 90,363 patients were admitted from the ICD-10 codes M16-M16.9. The CCG expenditure for osteoarthritis hip was estimated as £424m, which was the multiple of this ratio and the total expenditure for the whole PB category 15x (£4.9bn).

*Step 2. Estimate specialised services costs.*

Since the enactment of the Health and Social Care Act 2012, the NHS England budget for specialised services have been allocated nationally rather than locally. As a result, the related expenditure was no longer reported in the current PB dataset since 2013/14. Following Briggs et al. (2018), we obtained the last reported specialised service expenditure by individual disease from the 2012/13 PB dataset. For each relevant PB disease category, the expenditure on 'Other Secondary Care' in 2012/13 is divided by the total expenditure reported in the data. This relative ratio between specialised services costs and total expenditure is then multiplied by the disease-specific 2018/19 CCG expenditure calculated in step 1 to estimate the specialised services costs of modelled diseases in 2018/19. Using osteoarthritis hip as an example, the ratio of specialised service expenditure to total expenditure for the PB category *15x* in 2012/13 was 0.13. The estimated specialised services costs for osteoarthritis hip were thus £56m (£424m multiplied by 0.13).

For cancer subtypes, the majority of `Other Secondary Care' expenditure for chemotherapy and radiotherapy was allocated to category *02x*, *cancers and tumours*, *other* rather than to each specific cancer. The respective expenditure ratio of specialised services to total expenditure is therefore calculated based on the entirety of PB category *02*, *cancers and tumours* rather than the cancer subtype PB category. The specialised services costs for each modelled cancer subtype are then estimated by multiplying this ratio (0.34) with the total 2018/19 disease-specific CCG spend calculated in step 1.

*Step 3. Assign primary care costs.*

Apart from spending on prescribing, the PB dataset does not capture NHS England expenditure on primary care as these services are commissioned nationally. There is thus no data available on primary care expenditure by individual disease. To assign primary care costs to each modelled disease, the primary care prescribing expenditure reported in the 2018/19 PB data is used with the assumption that primary care expenditure on a given disease is proportional to the amount spent on primary care prescribing. For each PB category, the respective primary care prescribing expenditure is divided by the total primary care prescribing expenditure. The result is then multiplied by the total primary care expenditure in 2018/19 (£11.4bn) to obtain an estimate of primary care costs for each PB category. Disease-specific primary care costs are then obtained by multiplying this estimate with the admission ratio for each modelled disease calculated in step 1. For example, £298m was spent on primary care prescribing for PB category *15x* in 2018/19. This was divided by the total primary care prescribing expenditure reported in the PB data (£8,165m) and multiplied by the ratio of admission from PB category *15x* that was related to osteoarthritis hip computed in step 1. The result (0.003) gives the proportion of primary care expenditure spent on osteoarthritis hip. Multiplying this ratio with the total primary care spent in 2018/19, we obtained a primary care spent estimate of £36m on osteoarthritis hip in 2018/19.

*Step 4. Estimate NHS England expenditure on modelled and non-modelled diseases.*

For each disease, the CCG expenditure and costs spent on specialised services and primary care, computed in Steps 1 – 3, were added together to generate the total NHS England expenditure on the respective disease in 2018/19.

The NHS England costs on other diseases are the sum of CCG, primary care, specialised services expenditure that are not allocated to the diseases modelled in PRIMEtime and not allocated to programme budgeting categories *21*, *healthy individuals; 22*, *social care needs*; and *23*, *other*.

#### References

Department of Health and Social Care. DHSC annual report and accounts: 2018 to 2019. 11 Jul 2019. https://www.gov.uk/government/publications/dhsc-annual-report-and-accounts-2018-to-2019.

Briggs, A. D., Scarborough, P., & Wolstenholme, J. (2018). Estimating comparable English healthcare costs for multiple diseases and unrelated future costs for use in health and public health economic modelling. *PLoS One*, *13*(5), e0197257.

NHS Digital. Hospital Episode Statistics (HES) Database 2018/19 https://digital.nhs.uk/data-and-information/data-tools-and-services/data-services/hospital-episode-statistics

NHS England 2018-19 CCG Programme Budgeting Benchmarking Tool
