## Supplementary material for "PRIMEtime: an epidemiological model for informing diet and obesity policy": Text S3

### Text S3 – Additional results

*Table S1 Comparison of PRIMEtime estimates of disease prevalence for 2015 with estimates from GBD 2015*

| PRIMEtime | | GBD 2015 | |
|---|---|---|---|
| Disease | Prevalence (per 10,000) | Disease | Prevalence (per 10,000) |
| Breast cancer | 115.4 | Breast cancer | 83.3 |
| Ischemic heart disease | 291.6 | Ischemic heart disease | 278.3 |
| Ischemic stroke | 90.2 | Ischemic stroke | 85.9 |
| Atrial fibrillation and flutter | 186.0 | Atrial fibrillation and flutter | 191.1 |
| Diabetes mellitus type 2 | 881.4 | Diabetes mellitus type 2 | 1135.3 |
| Hypertensive heart disease | 26.1 | Hypertensive heart disease | 9.9 |
| Colon and rectum cancer | 48.4 | Colon and rectum cancer | 41.7 |
| Intracerebral haemorrhage | 15.0 | Intracerebral hemorrhage | 12.3 |
| Oesophageal cancer | 4.1 | Esophageal cancer | 2.5 |
| Pancreatic cancer | 2.7 | Pancreatic cancer | 1.3 |
| Multiple myeloma | 4.5 | Multiple myeloma | 3.5 |
| Kidney cancer | 8.7 | Kidney cancer | 7.7 |
| Liver cancer | 2.1 | Liver cancer | 1.5 |
| Uterine cancer | 11.6 | Uterine cancer | 11.4 |

*Table S2 Comparison of PRIMEtime estimates of disease prevalence for 2010-2015 with estimates from Kuan et al 2019*

| PRIMEtime | | Kuan et al 2019 | |
|---|---|---|---|
| Disease | Prevalence (per 10,000) | Disease | Prevalence (per 10,000) |
| Diabetes mellitus type 2 | 873.2 | Type 2 Diabetes Mellitus | 599.6 |
| Ischemic heart disease | 297.8 | Myocardial Infarction | 299 |
| Breast cancer | 117.1 | Primary Malignancy – Breast | 138.5 |
| Ischemic stroke | 93.9 | Ischaemic Stroke | 134.2 |
| Atrial fibrillation and flutter | 188.3 | Atrial Fibrillation | 432.9 |
| Liver cancer | 1.9 | Primary Malignancy – Liver | 4.3 |
| Multiple myeloma | 4.4 | Plasma Cell Malignancy | 10.4 |
| Pancreatic cancer | 2.7 | Primary Malignancy – Pancreas | 9.9 |
| Oesophageal cancer | 4.0 | Primary Malignancy – Oesophageal | 13 |
| Kidney cancer | 8.5 | Primary Malignancy – Kidney | 17.5 |
| Intracerebral haemorrhage | 15.6 | Intracerebral Haemorrhage | 31.5 |
| Uterine cancer | 11.2 | Primary Malignancy – Uterus | 36.9 |
| Colon and rectum cancer | 48.4 | Primary Malignancy – Bowel | 80 |

*Table S3 Change in prevalence (%) of healthy weight, overweight and obesity through time*

| | Male | | | Female | | |
|---|---|---|---|---|---|---|
| **Year** | **Healthy** | **Overweight** | **Obese** | **Healthy** | **Overweight** | **Obese** |
| **SSB tax** | | | | | | |
| 2015-2019 | 0.19 (0.058 to 0.31) | -0.061 (-0.1 to -0.019) | -0.12 (-0.21 to -0.039) | 0.2 (0.063 to 0.34) | -0.07 (-0.12 to -0.022) | -0.13 (-0.22 to -0.041) |
| 2020-2024 | 0.2 (0.063 to 0.35) | -0.061 (-0.1 to -0.019) | -0.14 (-0.24 to -0.044) | 0.22 (0.068 to 0.37) | -0.071 (-0.12 to -0.022) | -0.15 (-0.25 to -0.046) |
| 2025-2029 | 0.2 (0.061 to 0.33) | -0.058 (-0.099 to -0.018) | -0.14 (-0.23 to -0.043) | 0.21 (0.066 to 0.36) | -0.068 (-0.12 to -0.021) | -0.14 (-0.25 to -0.045) |
| 2030-2034 | 0.19 (0.058 to 0.32) | -0.056 (-0.096 to -0.018) | -0.13 (-0.22 to -0.041) | 0.2 (0.064 to 0.35) | -0.065 (-0.11 to -0.02) | -0.14 (-0.24 to -0.043) |
| 2035-2039 | 0.18 (0.057 to 0.31) | -0.055 (-0.094 to -0.017) | -0.13 (-0.21 to -0.039) | 0.2 (0.062 to 0.34) | -0.063 (-0.11 to -0.02) | -0.14 (-0.23 to -0.042) |
| 2040-2044 | 0.18 (0.057 to 0.31) | -0.056 (-0.095 to -0.017) | -0.13 (-0.21 to -0.039) | 0.2 (0.062 to 0.34) | -0.063 (-0.11 to -0.02) | -0.14 (-0.23 to -0.042) |
| 2045-2049 | 0.18 (0.057 to 0.31) | -0.057 (-0.097 to -0.018) | -0.13 (-0.21 to -0.039) | 0.2 (0.062 to 0.34) | -0.064 (-0.11 to -0.02) | -0.14 (-0.23 to -0.042) |
| 2050-2054 | 0.18 (0.057 to 0.31) | -0.058 (-0.098 to -0.018) | -0.13 (-0.21 to -0.039) | 0.2 (0.062 to 0.34) | -0.065 (-0.11 to -0.02) | -0.14 (-0.23 to -0.042) |
| **TV ad bans** | | | | | | |
| 2015-2019 | 0.31 (0.021 to 0.58) | -0.21 (-0.41 to -0.014) | -0.094 (-0.18 to -0.0064) | 0.34 (0.023 to 0.65) | -0.21 (-0.41 to -0.014) | -0.13 (-0.24 to -0.0088) |
| 2020-2024 | 0.36 (0.024 to 0.69) | -0.23 (-0.45 to -0.016) | -0.12 (-0.24 to -0.0085) | 0.41 (0.027 to 0.78) | -0.24 (-0.46 to -0.016) | -0.17 (-0.32 to -0.011) |
| 2025-2029 | 0.4 (0.027 to 0.77) | -0.24 (-0.47 to -0.016) | -0.16 (-0.3 to -0.011) | 0.46 (0.031 to 0.89) | -0.25 (-0.48 to -0.017) | -0.21 (-0.4 to -0.014) |
| 2030-2034 | 0.44 (0.03 to 0.85) | -0.25 (-0.47 to -0.016) | -0.2 (-0.37 to -0.013) | 0.51 (0.034 to 0.98) | -0.26 (-0.5 to -0.017) | -0.26 (-0.49 to -0.017) |
| 2035-2039 | 0.48 (0.032 to 0.92) | -0.25 (-0.47 to -0.016) | -0.23 (-0.44 to -0.016) | 0.56 (0.038 to 1.1) | -0.26 (-0.5 to -0.017) | -0.3 (-0.57 to -0.02) |
| 2040-2044 | 0.53 (0.035 to 1) | -0.25 (-0.48 to -0.017) | -0.27 (-0.52 to -0.018) | 0.62 (0.042 to 1.2) | -0.27 (-0.53 to -0.018) | -0.35 (-0.67 to -0.023) |
| 2045-2049 | 0.57 (0.038 to 1.1) | -0.26 (-0.5 to -0.017) | -0.32 (-0.61 to -0.021) | 0.69 (0.046 to 1.3) | -0.28 (-0.55 to -0.019) | -0.4 (-0.77 to -0.027) |
| 2050-2054 | 0.62 (0.041 to 1.2) | -0.26 (-0.51 to -0.017) | -0.36 (-0.69 to -0.024) | 0.75 (0.05 to 1.4) | -0.3 (-0.57 to -0.02) | -0.46 (-0.87 to -0.03) |
| **Weight loss program** | | | | | | |
| 2015-2019 | 0.022 (0.015 to 0.03) | 0.031 (0.021 to 0.041) | -0.053 (-0.071 to -0.035) | 0.027 (0.018 to 0.037) | 0.025 (0.017 to 0.033) | -0.052 (-0.07 to -0.035) |
| 2020-2024 | 0.017 (0.0096 to 0.025) | 0.022 (0.012 to 0.032) | -0.039 (-0.058 to -0.022) | 0.021 (0.012 to 0.03) | 0.018 (0.01 to 0.027) | -0.039 (-0.057 to -0.022) |
| 2025-2029 | 0.013 (0.0044 to 0.021) | 0.014 (0.0044 to 0.025) | -0.027 (-0.046 to -0.0088) | 0.015 (0.005 to 0.025) | 0.012 (0.0037 to 0.021) | -0.027 (-0.046 to -0.0088) |
| 2030-2034 | 0.0086 (0.002 to 0.018) | 0.0081 (0.0012 to 0.018) | -0.017 (-0.035 to -0.0032) | 0.0098 (0.0021 to 0.02) | 0.0069 (0.0011 to 0.015) | -0.017 (-0.036 to -0.0032) |
| 2035-2039 | 0.0059 (0.002 to 0.014) | 0.0046 (0.0012 to 0.013) | -0.01 (-0.027 to -0.0032) | 0.0065 (0.0021 to 0.016) | 0.0039 (0.001 to 0.011) | -0.01 (-0.027 to -0.0031) |
| 2040-2044 | 0.0048 (0.002 to 0.011) | 0.0034 (0.0012 to 0.0098) | -0.0082 (-0.021 to -0.0032) | 0.0052 (0.0021 to 0.013) | 0.0029 (0.001 to 0.0084) | -0.008 (-0.021 to -0.0031) |
| 2045-2049 | 0.0045 (0.002 to 0.0091) | 0.003 (0.0012 to 0.0073) | -0.0075 (-0.017 to -0.0031) | 0.0048 (0.0021 to 0.01) | 0.0025 (0.001 to 0.0061) | -0.0073 (-0.016 to -0.0031) |
| 2050-2054 | 0.0044 (0.002 to 0.008) | 0.0029 (0.0012 to 0.006) | -0.0074 (-0.014 to -0.0031) | 0.0047 (0.0021 to 0.0085) | 0.0025 (0.001 to 0.005) | -0.0071 (-0.013 to -0.0031) |
| **Combination** | | | | | | |
| 2015-2019 | 0.51 (0.2 to 0.82) | -0.24 (-0.44 to -0.04) | -0.27 (-0.39 to -0.15) | 0.57 (0.22 to 0.9) | -0.26 (-0.46 to -0.053) | -0.31 (-0.45 to -0.16) |
| 2020-2024 | 0.58 (0.22 to 0.93) | -0.27 (-0.49 to -0.051) | -0.31 (-0.45 to -0.15) | 0.64 (0.24 to 1) | -0.29 (-0.51 to -0.063) | -0.35 (-0.54 to -0.16) |
| 2025-2029 | 0.61 (0.21 to 1) | -0.29 (-0.52 to -0.058) | -0.32 (-0.5 to -0.15) | 0.69 (0.24 to 1.1) | -0.31 (-0.54 to -0.069) | -0.38 (-0.6 to -0.16) |
| 2030-2034 | 0.63 (0.2 to 1.1) | -0.29 (-0.52 to -0.063) | -0.34 (-0.54 to -0.14) | 0.72 (0.23 to 1.2) | -0.31 (-0.56 to -0.071) | -0.41 (-0.66 to -0.16) |
| 2035-2039 | 0.66 (0.2 to 1.1) | -0.3 (-0.53 to -0.066) | -0.37 (-0.59 to -0.14) | 0.76 (0.22 to 1.3) | -0.32 (-0.57 to -0.074) | -0.44 (-0.73 to -0.15) |
| 2040-2044 | 0.71 (0.2 to 1.2) | -0.3 (-0.54 to -0.068) | -0.41 (-0.67 to -0.14) | 0.82 (0.22 to 1.4) | -0.33 (-0.59 to -0.075) | -0.49 (-0.82 to -0.15) |
| 2045-2049 | 0.76 (0.21 to 1.3) | -0.31 (-0.55 to -0.069) | -0.45 (-0.75 to -0.14) | 0.89 (0.23 to 1.5) | -0.35 (-0.62 to -0.077) | -0.54 (-0.92 to -0.15) |
| 2050-2054 | 0.81 (0.21 to 1.4) | -0.32 (-0.56 to -0.07) | -0.49 (-0.83 to -0.14) | 0.95 (0.24 to 1.7) | -0.36 (-0.64 to -0.08) | -0.59 (-1 to -0.16) |

*Table S4 Cases of modelled obesity-related diseases averted in the first ten years*

| Disease | Male | Female |
|---|---|---|
| **SSB tax** | | |
| Ischemic heart disease | -1,400 (-2,400 to -430) | -620 (-1,100 to -190) |
| Ischemic stroke | -190 (-340 to -57) | -200 (-370 to -60) |
| Intracerebral hemorrhage | -100 (-190 to -27) | -110 (-210 to -28) |
| Hypertensive heart disease | -130 (-260 to -29) | -150 (-310 to -37) |
| Atrial fibrillation and flutter | -530 (-970 to -160) | -450 (-820 to -130) |
| Diabetes mellitus type 2 | -6,400 (-11,000 to -1,900) | -5,300 (-9,200 to -1,600) |
| Breast cancer | - | -85 (-150 to -25) |
| Colon and rectum cancer | -15 (-30 to -3.9) | -14 (-28 to -3.6) |
| Esophageal cancer | -39 (-73 to -12) | -22 (-41 to -6.7) |
| Kidney cancer | -18 (-34 to -5.4) | -13 (-25 to -3.9) |
| Liver cancer | -21 (-41 to -5.7) | -16 (-32 to -4.4) |
| Multiple myeloma | -5.4 (-9.6 to -1.7) | -4.8 (-8.5 to -1.5) |
| Pancreatic cancer | -9.2 (-18 to -2.4) | -11 (-22 to -2.9) |
| Uterine cancer | - | -72 (-120 to -22) |
| **TV ad bans** | | |
| Ischemic heart disease | 0.0001 (0 to 0.0002) | 0.0002 (0 to 0.0004) |
| Ischemic stroke | - | 0.0001 (0 to 0.0001) |
| Intracerebral hemorrhage | - | - |
| Hypertensive heart disease | - | - |
| Atrial fibrillation and flutter | -0.23 (-0.46 to -0.015) | -0.16 (-0.32 to -0.011) |
| Diabetes mellitus type 2 | 0.0047 (0.0003 to 0.0094) | 0.0039 (0.0003 to 0.0078) |
| Breast cancer | - | -3.8 (-7.7 to -0.28) |
| Colon and rectum cancer | -0.2 (-0.42 to -0.012) | -0.28 (-0.6 to -0.017) |
| Esophageal cancer | -0.11 (-0.24 to -0.0068) | -0.098 (-0.2 to -0.0059) |
| Kidney cancer | -0.58 (-1.2 to -0.035) | -0.66 (-1.4 to -0.04) |
| Liver cancer | -0.36 (-0.79 to -0.023) | -0.28 (-0.63 to -0.018) |
| Multiple myeloma | -0.018 (-0.036 to -0.0012) | -0.016 (-0.031 to -0.001) |
| Pancreatic cancer | -0.029 (-0.061 to -0.0018) | -0.04 (-0.085 to -0.0025) |
| Uterine cancer | - | -1.7 (-3.3 to -0.11) |
| **Weight loss program** | | |
| Ischemic heart disease | -390 (-560 to -230) | -180 (-270 to -110) |
| Ischemic stroke | -54 (-80 to -32) | -59 (-91 to -34) |
| Intracerebral hemorrhage | -29 (-46 to -14) | -31 (-51 to -14) |
| Hypertensive heart disease | -35 (-61 to -13) | -43 (-74 to -18) |
| Atrial fibrillation and flutter | -150 (-220 to -80) | -130 (-200 to -70) |
| Diabetes mellitus type 2 | -1,700 (-2,400 to -1,000) | -1,500 (-2,200 to -920) |
| Breast cancer | - | -26 (-38 to -15) |
| Colon and rectum cancer | -4.3 (-7.3 to -1.9) | -4.1 (-7 to -1.9) |
| Esophageal cancer | -12 (-19 to -7) | -7.1 (-11 to -4) |
| Kidney cancer | -5.5 (-8.4 to -3.1) | -4.1 (-6.3 to -2.3) |
| Liver cancer | -6.2 (-11 to -2.9) | -4.9 (-8.5 to -2.2) |
| Multiple myeloma | -1.6 (-2.4 to -0.93) | -1.4 (-2.2 to -0.85) |
| Pancreatic cancer | -2.7 (-4.5 to -1.3) | -3.3 (-5.5 to -1.6) |
| Uterine cancer | - | -23 (-31 to -14) |
| **Combination** | | |
| Ischemic heart disease | -1,800 (-2,800 to -800) | -800 (-1,300 to -360) |
| Ischemic stroke | -240 (-400 to -110) | -260 (-440 to -110) |
| Intracerebral hemorrhage | -130 (-230 to -50) | -140 (-250 to -51) |
| Hypertensive heart disease | -160 (-310 to -50) | -190 (-360 to -66) |
| Atrial fibrillation and flutter | -670 (-1,100 to -290) | -570 (-980 to -250) |
| Diabetes mellitus type 2 | -8,100 (-13,000 to -3,500) | -6,800 (-11,000 to -3,100) |
| Breast cancer | - | -110 (-190 to -52) |
| Colon and rectum cancer | -20 (-36 to -7.3) | -19 (-34 to -6.9) |
| Esophageal cancer | -52 (-87 to -23) | -29 (-49 to -13) |
| Kidney cancer | -24 (-41 to -11) | -18 (-30 to -8.1) |
| Liver cancer | -28 (-50 to -11) | -21 (-39 to -8.1) |
| Multiple myeloma | -7 (-11 to -3.2) | -6.3 (-10 to -2.8) |
| Pancreatic cancer | -12 (-21 to -4.5) | -14 (-26 to -5.4) |
| Uterine cancer | - | -97 (-150 to -47) |

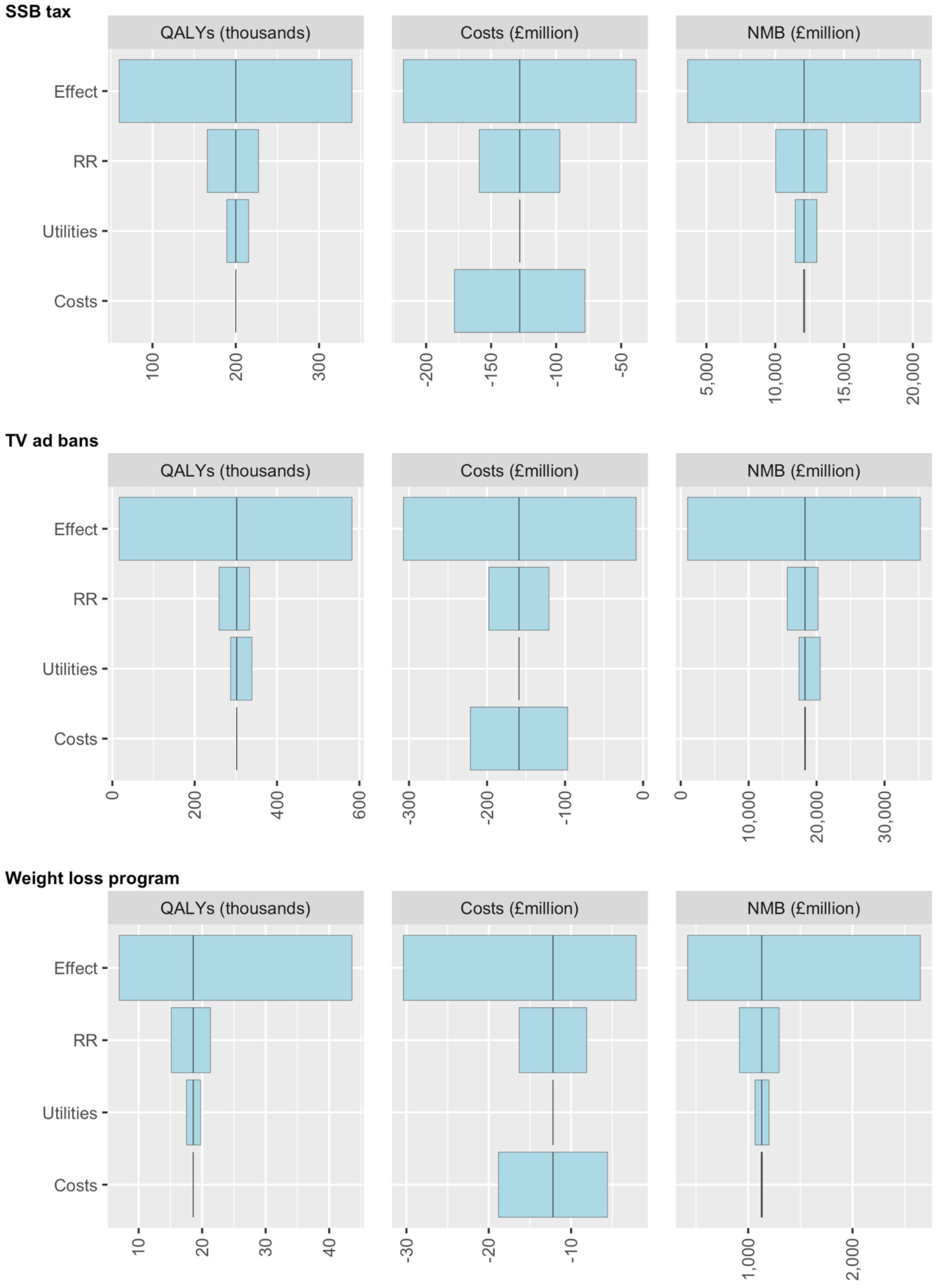


*Figure S1 Tornado diagrams showing relative impact of uncertainty around model inputs on predicted QALYs, costs and net monetary benefit (NMB), for the individual intervention scenarios*
